# Data Reduction in SARS-CoV-2 Amplicon Sequencing: Effects of Read Subsampling and Normalization on Consensus Quality, Lineage Assignment, and Computational Burden

**DOI:** 10.64898/2026.10.05.26363801

**Authors:** Yered Ortega, Pooja Gupta, Olinto Olinares-Perdomo, Kelly F. Oakeson, Erin L. Young

## Abstract

**Background:** Amplicon-based next-generation sequencing is a cornerstone of viral genomic surveillance, but it often produces datasets with highly uneven read coverage due to PCR amplification bias. This leads to excessive data generation and substantial computational overhead for downstream bioinformatic analysis.

**Results:** We evaluated the efficacy of two data reduction strategies on 220 SARS-CoV-2 clinical specimens and 12 wastewater samples: random subsampling with SeqKit and read depth normalization with BBNorm. FASTQ files were reduced to target depths from 10X to 10,000X to compare genome coverage, lineage assignment concordance, and computational resource usage. Both strategies markedly reduced total processing time. For clinical isolates, read depth normalization corrected for amplification bias resulting in highly concordant lineage assignments at a target depth of just 200X. Random subsampling preserved inherent bias but produced reliable assignments at depths over 500X. Reducing target depths below 500X in complex wastewater mixtures severely degraded variant deconvolution accuracy and heavily skewed lineage abundance profiles for both methods.

**Conclusion:** Both random subsampling and read depth normalization successfully reduce computational overhead while preserving key clinical sequencing metrics at appropriate reduction settings. The optimal method depends on the analytical goal, ranging from computationally lightweight random subsampling for rapid lineage assignment, to read depth normalization for generating high-quality consensus sequences. However, aggressive data reduction should be avoided in environmental surveillance, where maintaining a high baseline sequencing depth is critical for accurate mixed-variant deconvolution.

## Background

Next-generation sequencing (NGS) has become an invaluable tool for the genomic surveillance of infectious diseases, facilitating the monitoring of viral evolution and the emergence of variants of concern.^1,2^ Among the various NGS methods, amplicon-based sequencing has been widely adopted for pathogens like SARS-CoV-2, Zika, and Ebola, making it ideal for large-scale pathogen surveillance.^3–5^ A prominent example of this approach has been the ARTIC network’s protocol, which utilizes multiplexed, tiled polymerase chain reaction (PCR) amplicons to cover the majority of a viral genome.^3^

Amplicon-based sequencing is susceptible to technical biases that can distort the final data. The PCR process itself does not amplify all templates with equal efficiency.^6^ Factors such as amplicon length and GC content can cause certain sequences to be systematically over- or under-represented in the final library.^7,8^

In practice, this variability in amplification efficiency leads to highly uneven read depth across a reference genome.^6,9,10^ To compensate, sequencing runs are often performed far in excess of their theoretical need to ensure all genomic regions meeting minimum depth thresholds.^11^ For example, the SARS-CoV-2 genome is approximately 30,000 base pairs in length.^12^ For a 151 base-pair sequencing run, a theoretical minimum of only 20,000 reads would be required to achieve 100X average coverage, a depth sufficient for robust consensus generation and variant calling. This over-sequencing dramatically inflates downstream computational overhead, leading to prolonged analytical turnaround times, and increased processing costs.

Furthermore, viral genomic tracking has expanded beyond single-isolate clinical samples to include complex environmental matrices like wastewater.^13–15^ Wastewater samples contain highly fragmented, mixed viral populations, requiring specialized deconvolution algorithms to estimate the relative abundance of closely related subvariants. Accurate deconvolution is uniquely sensitive to sequencing depth and coverage evenness, as algorithms must reliably detect linked minority mutations across the entire genome to successfully unmix lineages.

To mitigate the computational burden of amplicon datasets, genomic monitoring programs can implement data reduction steps prior to downstream analysis. This study evaluates two distinct computational strategies: random subsampling (which proportionately reduces reads, inherently preserving the original PCR bias) and read depth normalization (which algorithmically targets bias by downsampling over-represented sequence regions and capping reads from over-represented amplicons). We assessed these two methods on 220 SARS-CoV-2 clinical samples and 12 wastewater specimens to determine their impact on genome completeness, Pango lineage assignment concordance, mixed-variant deconvolution, and overall pipeline efficiency.

## Methods

### Sample Processing and Sequencing

Clinical SARS-CoV-2 positive specimens (n=220) and environmental wastewater samples (n=12) were processed for whole-genome sequencing using Illumina’s COVIDSeq protocol with ARTIC v5.3.2 primers, according to the manufacturer’s instructions. The resulting amplicon pools were sequenced on an Illumina NextSeq 2000 platform to generate paired-end short-read data, which were subsequently submitted to the Sequence Read Archive (SRA).^16^ A list of SRA accessions for samples is found in Supplemental Table 1.

### Bioinformatic Processing

Raw FASTQ files were downloaded directly from the SRA using fasterq-dump.^16,17^ To isolate computational overhead, file download times were excluded from benchmarking. All benchmark runs were performed on a dedicated Linux workstation equipped with an Intel Xeon w9-3495X processor (56 physical cores, 112 logical threads), 512 GB RAM, and a local 3.8 TB NVMe SSD. No other user jobs or background processes were active during testing.

Benchmarks were run sequentially. Each sample configuration was executed as a single run without repeated replicates. Processing wall-clock time, peak memory (maximum resident set size, %M) and CPU utilization (%P) were measured using GNU time (v1.7). Average processing time reflects the mean elapsed wall-clock time across all target depth conditions.

Raw reads were reduced using either read depth normalization with BBNorm (v39.33) or random subsampling with SeqKit (v2.8.2) to target depths ranging from 10X to 10,000X. Because SeqKit lacks a parameter for specific coverage targets, the required read counts were calculated using a 151 base-pair read length and the reference genome size (Supplemental Table 2). Because BBNorm and SeqKit produced different realized mean depths at equivalent nominal targets, comparisons should be interpreted as comparisons of the two reduction workflows at their respective operating settings rather than as strictly depth-matched comparisons. Following reduction, reads underwent quality filtering with FASTP (v1.0.1).^18–20^

### Alignment and Consensus Generation

All read subsets were aligned to the Wuhan-Hu-1 reference genome (GenBank: NC_045512.2) using BBMap (v39.33).^19^ Primer sequences were trimmed from the resulting BAM files using iVar (v1.4.4), which was also utilized to generate consensus sequences applying a minimum depth threshold of 10X and a majority frequency threshold of 0.5.^21^ SAMtools (v1.22.1) was used for sorting and indexing alignment files.^22^

### Data Comparison

The resulting FASTA and SAM/BAM files from each pipeline were used to calculate and compare key sequencing metrics. Genome coverage, number of covered bases, mean base quality, mean mapping quality, and mean read depth were obtained directly from the output of SAMtools coverage (v1.22.1).^22,23^ The column names and metric definitions reported in Table 2 follow the samtools coverage output format. Metrics for each amplicon at each processing step and for each sample were assessed using SAMtools ampliconstats (v1.22.1).^22,23^ Pango lineages were determined using Pangolin (v4.3.1), ^24^ and parental lineages were assigned using pango_aliasor (v0.3.0) to evaluate classification specificity.^25^ Wastewater fine-scaled mixed lineage populations were deconvoluted and assigned using Freyja (v2.0.3).^13^

## Results

### Overview of the SARS-CoV-2 Genomic Dataset

A total of 220 SARS-CoV-2 positive clinical specimens and 12 wastewater samples were collected in Utah between December 2023 and April 2025 as part of baseline genomic surveillance (Table 1; Supplemental Table 1). Sequencing produced highly variable data volumes, averaging 16.4 million reads per sample (range: 104.6 thousand to 140.5 million reads). All clinical isolates were identified as Omicron variants, encompassing 78 distinct Pango lineages (Supplemental Table 3). This unprocessed dataset served as the baseline to evaluate the performance of the data reduction strategies.

**Table 1.** SARS-CoV-2 Genomic Dataset Overview.

| Metric | Value |
| --- | --- |
| **Sample Characteristics** |  |
| Number of Samples | 220 |
| **Sequencing Metrics** |  |
| Average Number of Reads per Sample | 16,378,884 |
| Range of Reads per Sample | 104,692 – 140,466,898 |
| Average Expected Coverage | 82,440X |
| **Genomic Characteristics** |  |
| SCORPIO Call: Omicron | 196 |
| SCORPIO Call: Probable Omicron | 18 |
| SCORPIO Call: No call | 6 |
| Total PANGO Lineages Observed | 78 |
| Top 3 Lineages (*n* samples) | XEC (26), LP.8.1 (23), LP.8.1.1 (21) |

### Effect of Data Reduction on Genome Completeness

To assess the impact of data reduction on assembly quality, we aligned each read subset to the reference genome and compared key metrics (Figure 1; Table 2; Supplemental Tables 3, 5). The principal effect of reduction was on genome completeness; base quality was essentially unchanged, while mapping quality showed a modest method-dependent effect. Read depth normalization with BBNorm consistently yielded higher median genome coverage and more covered bases than random subsampling with SeqKit, particularly at lower depths (Figure 1A–B). At the tested operating settings, BBNorm produced highly complete genomes at its 200X nominal setting, whereas SeqKit required a nominal setting above 500X to achieve comparable completeness. Notably, minor method-dependent artifacts emerged: BBNorm induced a slight decrease and increased variance in mean mapping quality, whereas SeqKit maintained a stable quality profile across all depths (Figure 1C–D).

**Figure 1.**
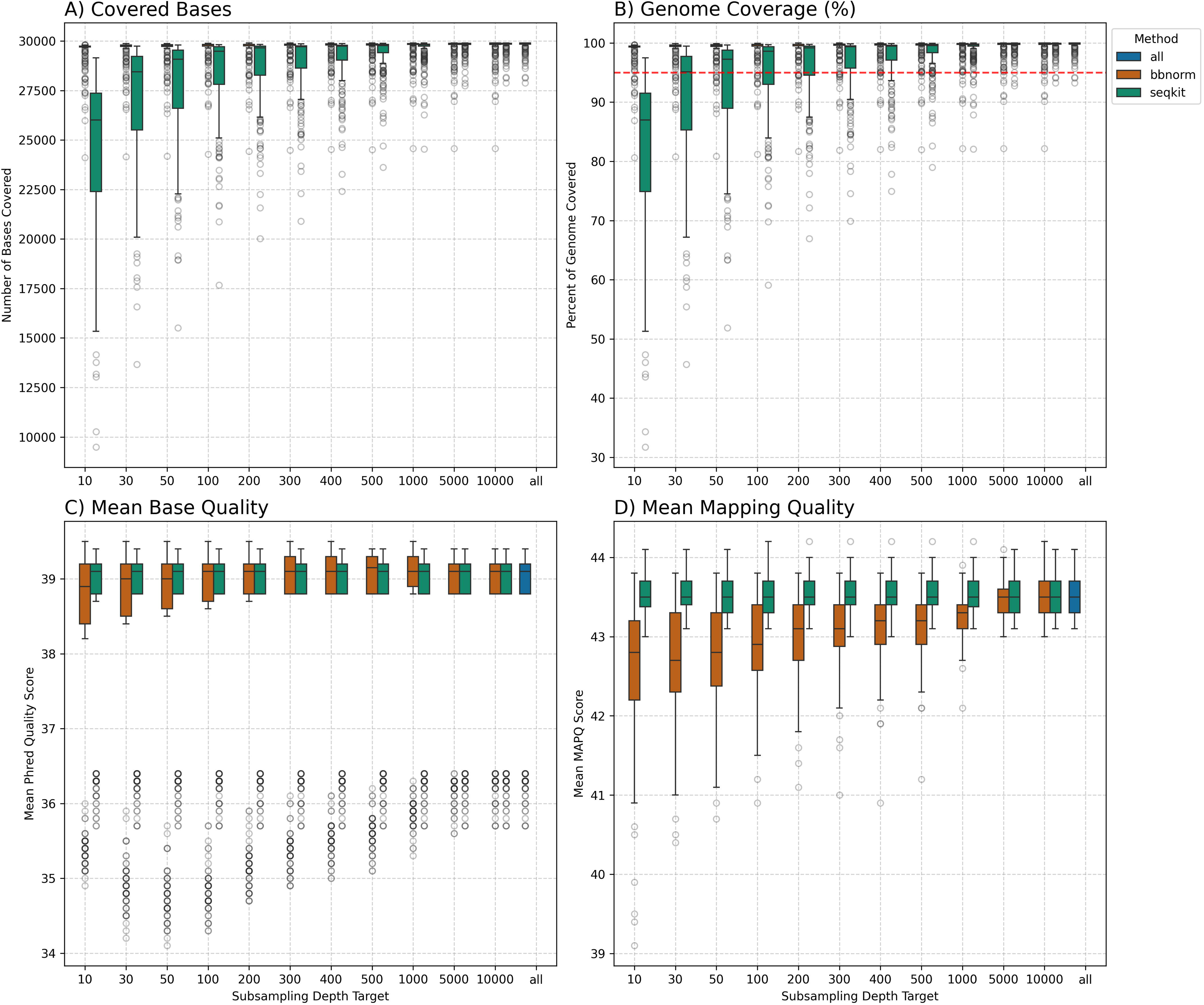
Effect of Data Reduction on Genome-Wide Quality Metrics. Boxplots compare key genome quality metrics from the full, unprocessed dataset (blue) to datasets reduced by either read depth normalization with BBNorm (orange) or random subsampling with SeqKit (green). Each boxplot represents the distribution of values across all 220 samples at the indicated target depth. The panels show: (A) the total number of covered bases, (B) the percentage of the genome covered with a dashed red line denoting 95% genome coverage, (C) the mean base quality, and (D) the mean mapping quality of mapped reads.

**Table 2.**
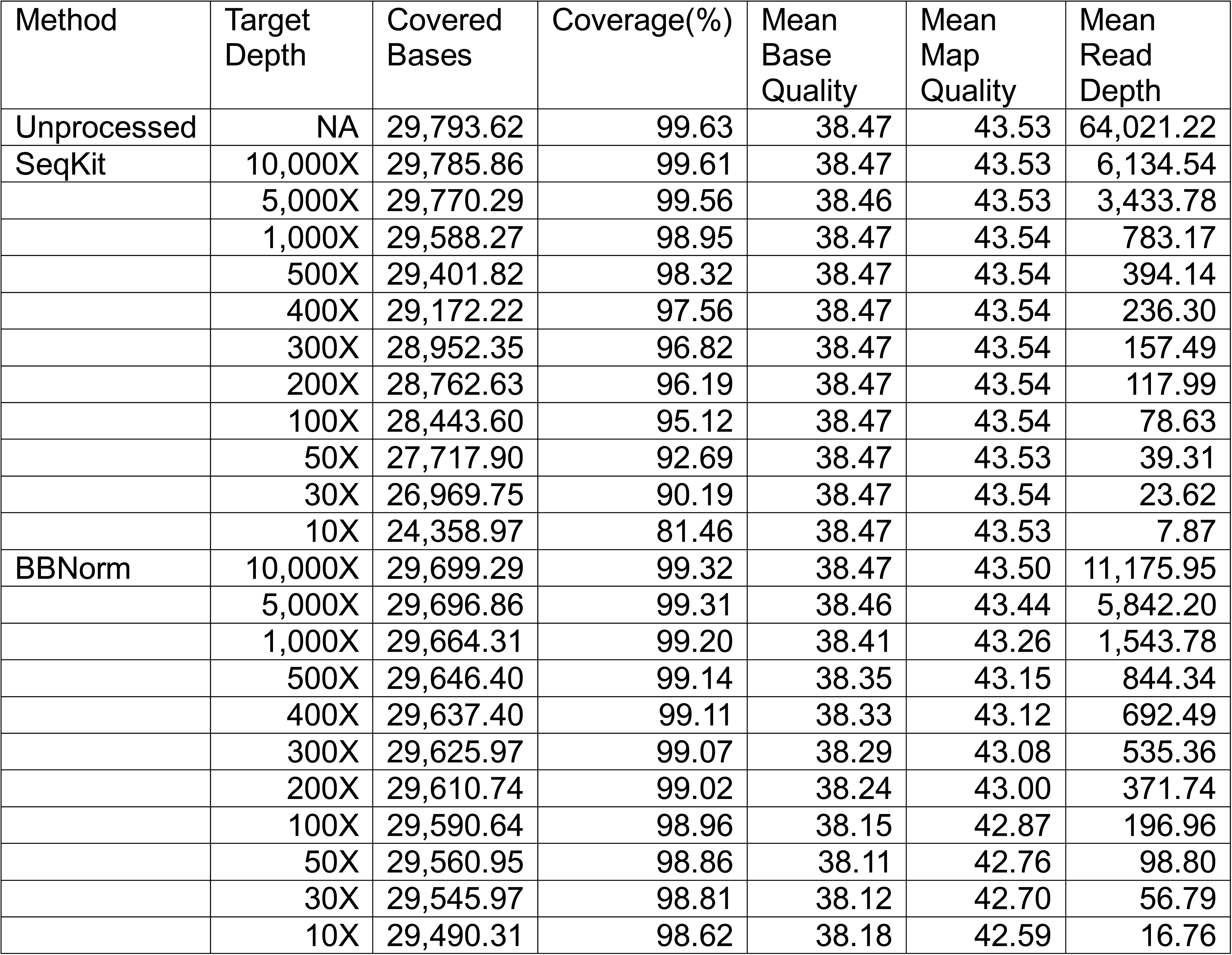
Genome coverage and sequencing quality metrics following read reduction.

### Impact on Amplification Bias

Analyzing read distribution at the individual amplicon level revealed how each method interacts with multiplex PCR amplification bias (Figure 2; Supplemental Tables 6–9). Unprocessed datasets exhibited severe bias, with specific amplicons systematically over- or under-represented (Figure 2A). SeqKit proportionally reduced reads, strictly preserving this uneven distribution. Conversely, BBNorm actively equalized read distribution, generating highly uniform coverage across the amplicon scheme (Figure 2A–C). Consequently, fewer amplicons failed quality control (<10X depth) in the normalized datasets compared to the subsampled datasets at every target depth (Figure 2D).

**Figure 2.**
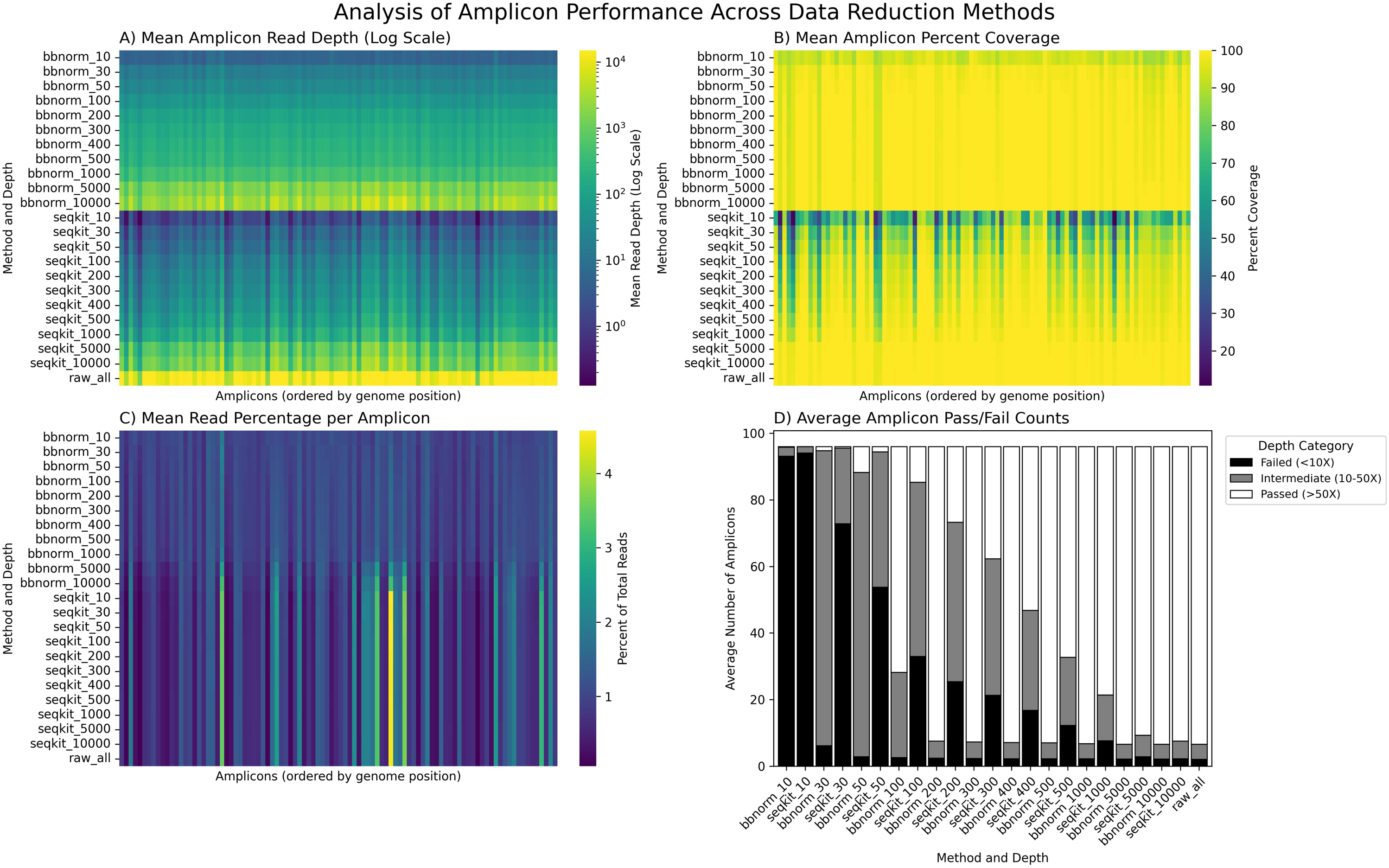
Read Depth Normalization Corrects for Amplicon-Level PCR Bias. This figure illustrates how read depth normalization (BBNorm) and random subsampling (SeqKit) impact read distribution across the SARS-CoV-2 genome amplicons. The heatmaps for (A) mean read depth, (B) percent coverage, and (C) percentage of total reads show the performance for each amplicon (columns) under each condition (rows). The more uniform BBNorm profiles are consistent with reduced representation of amplification-driven coverage bias. (D) The stacked bar chart shows the average number of amplicons per sample that met depth-based quality control thresholds, categorized as “Passed” (>50X depth), intermediate (10X−50X), or “Failed” (<10X).

### Influence on Consensus Quality and Lineage Assignment

Differences in genome completeness directly impacted downstream Pango lineage assignment (Figure 3; Supplemental Table 4). Consensus sequences derived from normalized data contained markedly fewer ambiguous bases (‘N’s) than those from subsampled data at matched depths (Figure 3A). This translated to superior classification stability: BBNorm provided lineage assignments concordant down to a 200X target depth (Figure 3C). In contrast, SeqKit datasets suffered higher classification failure and mismatch rates at depths below 500X (Figure 3D).

**Figure 3.**
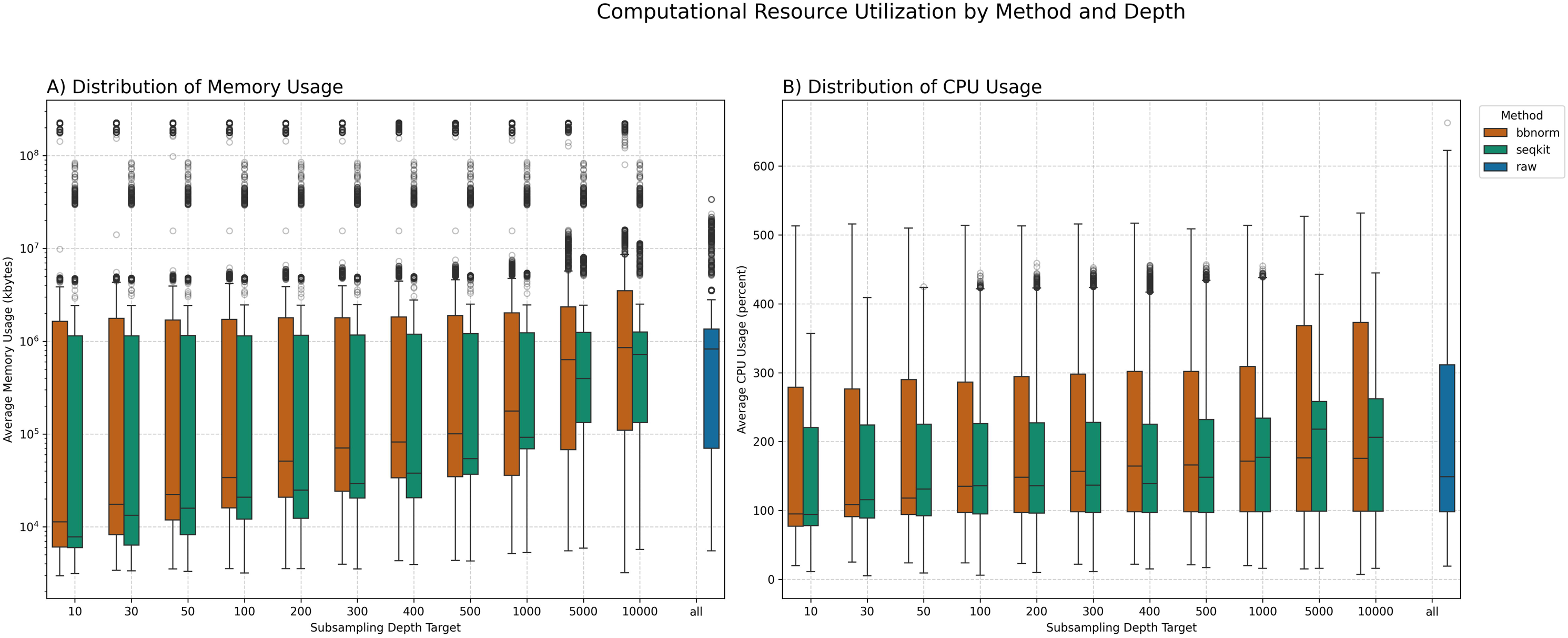
Impact of Data Reduction on Consensus Quality and Lineage Assignment. The figure evaluates how data reduction affects consensus sequence quality and Pango lineage assignment concordance. The top row compares quality metrics for datasets reduced with BBNorm (orange) and SeqKit (green) against the full, unprocessed ‘raw’ dataset (blue). (A) Boxplots of ambiguity content show the percentage of ‘N’ bases in each consensus sequence. (B) Boxplots of Scorpio Support show Pangolin’s confidence score for each lineage call. The bottom row visualizes the concordance of lineage assignments for (C) BBNorm and (D) SeqKit. Each stacked bar shows the number of samples classified as a ‘Match’, ‘Mismatch’, or ‘Failure’ when compared to the original, full dataset.

### Computational Workload

Both data reduction strategies successfully decreased total pipeline processing time relative to the full, unprocessed datasets (Figure 4A; Supplemental Figures 1–3; Supplemental Table 10). While SeqKit acts as a computationally lightweight filter with minimal initial overhead, BBNorm requires a more resource-intensive, two-pass normalization step. However, processing fewer downstream reads ultimately yielded a net reduction in total time for both methods. Resource utilization followed a similar pattern, with reduced datasets generally requiring less memory and CPU than the unprocessed dataset (Figure 5).

**Figure 4.**
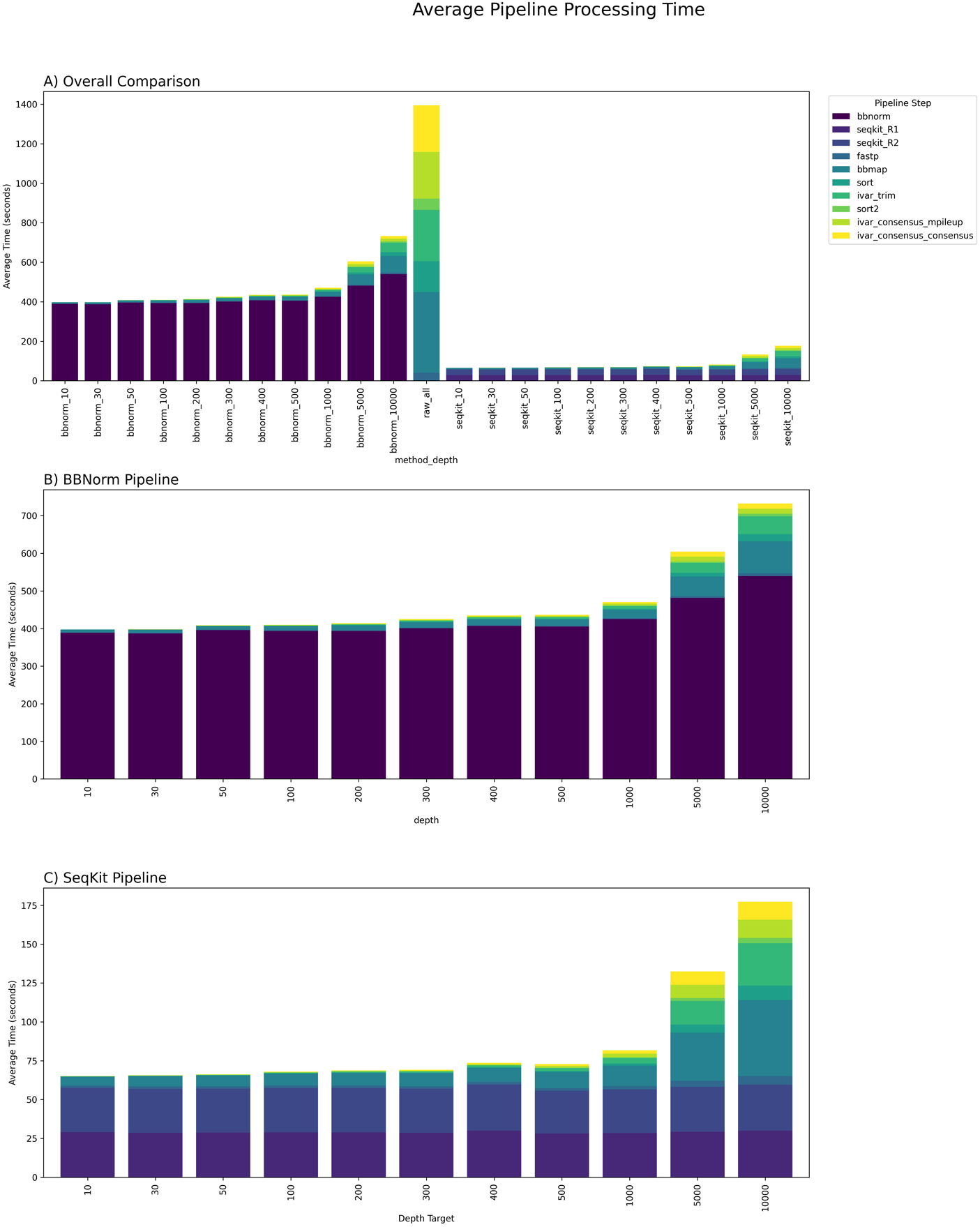
Average Pipeline Processing Time by Method. The figure presents the average processing time for the bioinformatic pipeline, broken down by individual steps. (A) The top panel provides a comprehensive overview comparing the unprocessed (‘raw’) dataset to all BBNorm and SeqKit datasets. (B) and (C) offer detailed views of the processing time for only the BBNorm and SeqKit pipelines, respectively. In all panels, the total height of each bar represents the total processing time, and each colored segment corresponds to the time taken by a specific pipeline step, as defined in the legend.

**Figure 5.**
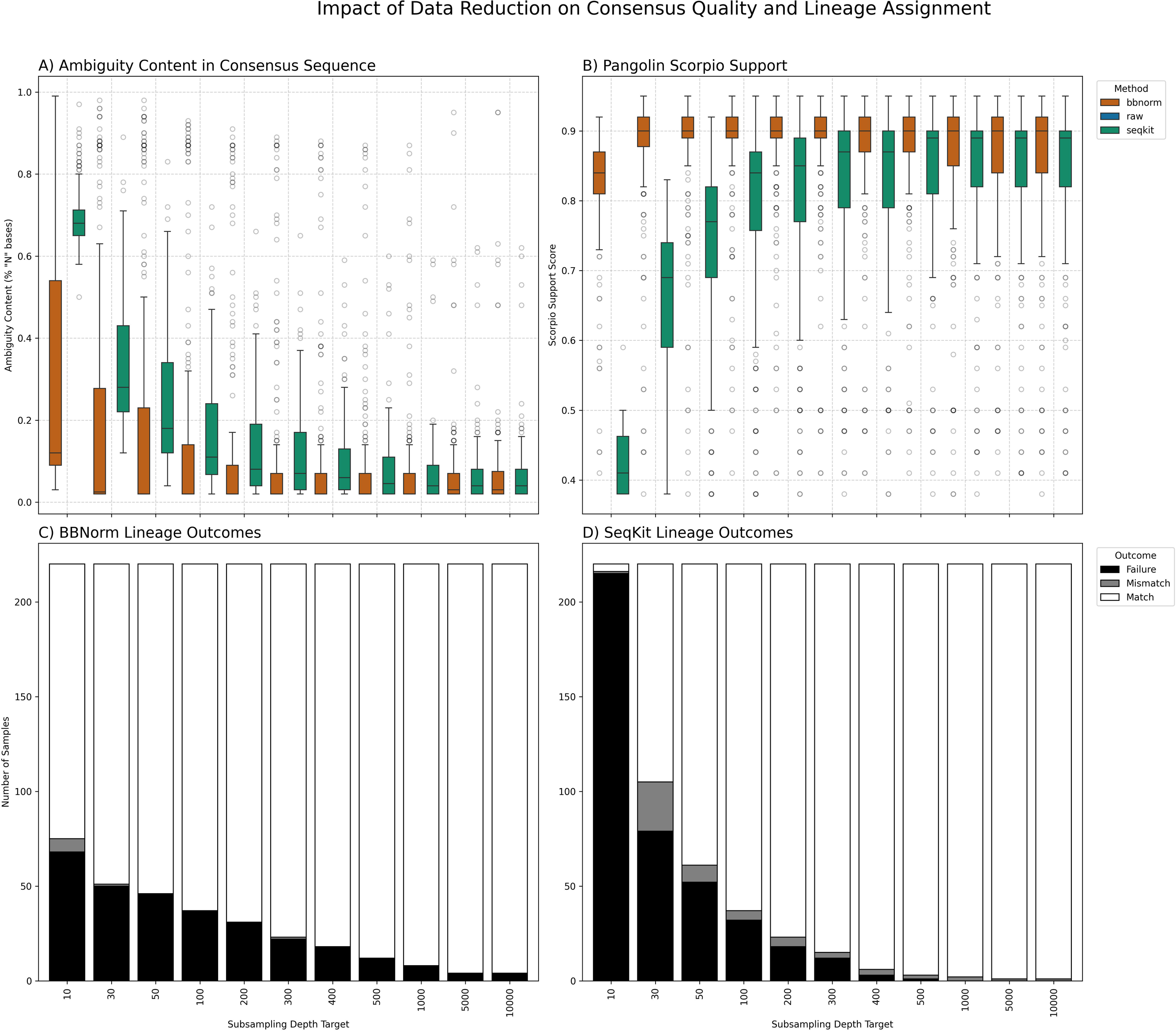
Computational Resource Utilization by Method and Depth. The figure shows the distribution of computational resource usage for the different data reduction strategies across all 220 samples. (A) Boxplots show the distribution of average memory usage (in kilobytes) on a logarithmic scale. (B) Boxplots show the distribution of average CPU usage (in percent). In both panels, results from the unprocessed ‘raw’ dataset (blue) are compared to those from datasets reduced by BBNorm (orange) and SeqKit (green).

### Mixed Lineage Impact

Accurate deconvolution of mixed viral populations in wastewater is highly sensitive to sequencing depth.^13–15^ We evaluated this by analyzing 12 wastewater samples (Table 3) subjected to progressively lower target depths (Table 4; Supplemental Table 11).

**Table 3.**
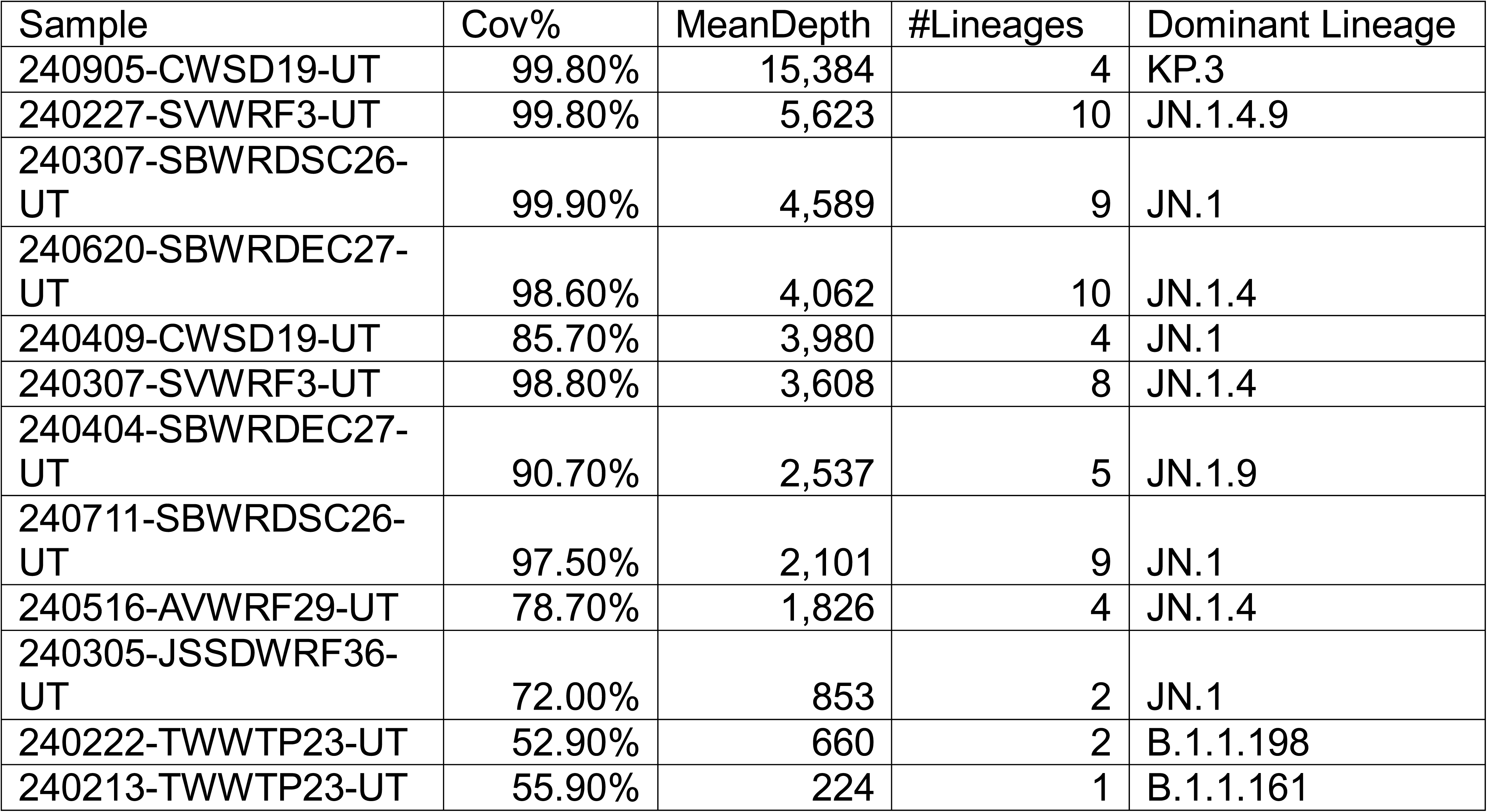
Wastewater Baseline Characteristics.

| Sample | Cov% | MeanDepth | #Lineages | Dominant Lineage |
| --- | --- | --- | --- | --- |
| 240905-CWSD19-UT | 99.80% | 15,384 | 4 | KP.3 |
| 240227-SVWRF3-UT | 99.80% | 5,623 | 10 | JN.1.4.9 |
| 240307-SBWRDSC26-UT | 99.90% | 4,589 | 9 | JN.1 |
| 240620-SBWRDEC27-UT | 98.60% | 4,062 | 10 | JN.1.4 |
| 240409-CWSD19-UT | 85.70% | 3,980 | 4 | JN.1 |
| 240307-SVWRF3-UT | 98.80% | 3,608 | 8 | JN.1.4 |
| 240404-SBWRDEC27-UT | 90.70% | 2,537 | 5 | JN.1.9 |
| 240711-SBWRDSC26-UT | 97.50% | 2,101 | 9 | JN.1 |
| 240516-AVWRF29-UT | 78.70% | 1,826 | 4 | JN.1.4 |
| 240305-JSSDWRF36-UT | 72.00% | 853 | 2 | JN.1 |
| 240222-TWWTP23-UT | 52.90% | 660 | 2 | B.1.1.198 |
| 240213-TWWTP23-UT | 55.90% | 224 | 1 | B.1.1.161 |

**Table 4.** Wastewater Lineage Shifts with Depth.

| Method | Target Depth | Avg. Lineages Detected | Original Dominant Abundance | Dominant Match Rate |
| --- | --- | --- | --- | --- |
| Unprocessed | NA | 8.6 | 56.10% | 100% |
| SeqKit | 10,000X | 8.3 | 44.30% | 83% |
|  | 5,000X | 7.9 | 53.40% | 100% |
|  | 1,000X | 8.2 | 44.30% | 83% |
|  | 500X | 7.6 | 52.00% | 92% |
|  | 400X | 7.9 | 44.90% | 83% |
|  | 300X | 7.1 | 45.10% | 67% |
|  | 200X | 8.2 | 40.30% | 75% |
|  | 100X | 5.9 | 45.50% | 67% |
|  | 50X | 7.8 | 42.40% | 83% |
|  | 30X | 5.2 | 44.80% | 67% |
|  | 10X | 7.2 | 41.60% | 67% |
| BBNorm | 10,000X | 5.4 | 43.90% | 67% |
|  | 5,000X | 6.1 | 38.60% | 67% |
|  | 1,000X | 5.1 | 35.80% | 50% |
|  | 500X | 6.7 | 47.00% | 58% |
|  | 400X | 6.7 | 34.90% | 50% |
|  | 300X | 5.9 | 47.50% | 67% |
|  | 200X | 7.5 | 31.70% | 50% |
|  | 100X | 5.2 | 47.40% | 58% |
|  | 50X | 9.6 | 16.10% | 25% |
|  | 30X | 4.2 | 40.80% | 42% |
|  | 10X | 97 | 13.70% | 27% |

SeqKit generally maintained higher concordance with the baseline dominant lineage at higher target depths, whereas BBNorm showed substantial variability across samples (Table 4). However, as depth decreased below 500X, Freyja increasingly failed to link defining subvariant mutations. By 100X, the true dominant lineage’s abundance dropped from a baseline average of 56.1% to 45.5% (SeqKit) and 47.4% (BBNorm).

Extreme data starvation (≤30X) induced profound deconvolution failures. At 10X, BBNorm exhibited severe spurious lineage inflation, artificially inflating the average number of detected lineages to 97.0 as sparse reads misaligned to obsolete basal lineages. SeqKit resisted this specific inflation, but its dominant lineage match rate fell to an unreliable 27%. Overall, aggressively subsampling complex environmental mixtures, particularly those with suboptimal baseline coverage, artificially can substantially distort inferred lineage diversity and composition.

## Discussion

Our findings demonstrate that both random subsampling (SeqKit) and read depth normalization (BBNorm) can substantially reduce the computational overhead of processing amplicon sequencing data, but the choice of data-reduction strategy matters because reducing reads and normalizing reads are not equivalent operations. Implementing a data reduction step drastically lowers the computational burden for bioinformatic analysis. These findings could inform future optimization of sequencing depth and sample multiplexing.

The primary challenge in amplicon-based sequencing is not the total volume of data, but its uneven distribution. Amplification bias, a known artifact of multiplex PCR, concentrates reads onto a high-efficiency subset of amplicons, leaving other regions under-sequenced.^21,26^ Our analysis confirms that random subsampling proportionally scales down the dataset, which inherently preserves the original, uneven read distribution of the sequencing run. In contrast, read depth normalization algorithmically mitigates this bias by downsampling over-represented sequence regions from over-represented amplicons, resulting in a more uniform coverage profile.

The optimal data reduction method relies entirely on the specific analytical goal. If the primary objective is rapid Pango lineage assignment, random subsampling is a highly efficient, lightweight tool that provides reliable quality at a target depth of 500X. However, for applications requiring high-quality, complete consensus sequences, read depth normalization is highly advantageous, as it provides robust genomes at a lower threshold of 200X and minimizes ambiguity.

A strategy optimized for single-consensus clinical sequencing should not automatically be applied to mixed-population sequencing. At higher target depths, inferred lineage profiles progressively approached the baseline lineage profiles. BBNorm better preserved lineage detectability under aggressive reduction, whereas random subsampling more closely reproduced baseline sublineage composition once sufficient depth was retained. Below approximately 500X, both methods showed substantial loss of fine-scale lineage resolution. These differences were consistent with amplicon-level coverage patterns: BBNorm largely preserved amplicon representation across target depths, whereas random subsampling produced progressively greater amplicon dropout as depth decreased.

Crucially, these recommendations apply specifically to single-consensus workflows. For applications designed to detect low-frequency intra-host variants (iSNVs) or unmix clinical co-infections, normalization is generally not recommended, as critical minority variant data could be algorithmically capped and lost.^26–28^

Furthermore, it is important to contextualize these performance metrics within the specific parameters of this study. The analyzed dataset was downloaded from the Sequence Read Archive (SRA) and consisted strictly of viral reads, as human host data had already been removed. In a live genomic surveillance pipeline, host read depletion represents a significant computational bottleneck. Additionally, these results were generated using SARS-CoV-2 data from the ARTIC v5.3.2 primer scheme sequenced on an Illumina platform. While the overarching principle of mitigating PCR bias through data reduction is broadly applicable, the specific computational time savings and optimal depth thresholds may vary for alternative sequencing technologies, different amplicon tiling strategies, or other viral pathogens. Since small changes in bioinformatic workflows can impact identified mutations, future work should aim to validate these thresholds across a wider array of targets and platforms.^29,30^

## Conclusion

This work demonstrates the significant practical benefits of incorporating a data reduction step into amplicon sequencing pipelines. Both random subsampling and read depth normalization successfully reduce computational overhead while maintaining analytical integrity, provided the chosen method aligns with the specific analytical goal. Random subsampling offers a rapid, lightweight solution for routine clinical lineage assignment, whereas read depth normalization yields the high-quality consensus sequences required for robust phylogenetic tracking. For the wastewater samples evaluated here, aggressive reduction below approximately 500X compromised fine-scale mixed lineage deconvolution, indicating that substantially higher sequencing depths may be necessary for reliable environmental surveillance. By strategically implementing these data reduction techniques where appropriate, the genomic surveillance community can build more sustainable bioinformatic pipelines that maximize the epidemiological and scientific value.

## Data Availability

All raw FASTQ files are publicly available through the Sequence Read Archive (SRA), with individual accession numbers detailed in Supplemental Table 1. Comprehensive datasets generated and analyzed during this study are provided in the supplementary tables. This includes dataset overviews, target read depths, alignment and coverage metrics (via SAMtools and BBMap), amplicon performance statistics, computational resource utilization per pipeline step, and lineage assignments (via Pangolin). Furthermore, all customized scripts and example commands used to execute the analytical workflows and generate figures are openly accessible at https://github.com/erinyoung/normalization_subsampling_scripts_and_data.

## Declarations

### Ethics approval and consent to participate

All viral genomic datasets analyzed in this study were de-identified prior to analysis and retrieved from the publicly available NCBI Sequence Read Archive (SRA; https://www.ncbi.nlm.nih.gov/sra). Individual SRA accession numbers are detailed in Supplemental Table 1. No human patient data, personal health information, or identifying clinical records were collected or accessed. Because this study relied solely on de-identified, publicly accessible sequence data, dedicated institutional ethics oversight was not required.

## Authors’ contributions

Yered Ortega: Curated clinical samples, designed the analytical methodology, executed the bioinformatic analysis, and contributed to manuscript preparation.

Pooja Gupta: Curated wastewater isolates and performed preliminary data processing and analysis.

Olinto Olinares-Perdomo: Initiated early analytical workflows and managed data curation, including FASTQ submission to the Sequence Read Archive (SRA).

Kelly F. Oakeson: Supervised sample acquisition and data submission processes.

Erin L. Young: Developed the custom scripts for data analysis, supervised the bioinformatic pipeline workflows, and led manuscript preparation.

## Acknowledgements

The authors gratefully acknowledge the dedicated laboratory and epidemiology staff at the Unified Public Health Laboratory (UPHL) and the State of Utah Department of Health and Human Services. Their extensive efforts in sample collection, processing, and ongoing epidemiological surveillance were essential in enabling the testing and data generation that made this analysis possible.

