## Supplemental Figure 4 for "Data Reduction in SARS-CoV-2 Amplicon Sequencing: Effects of Read Subsampling and Normalization on Consensus Quality, Lineage Assignment, and Computational Burden"

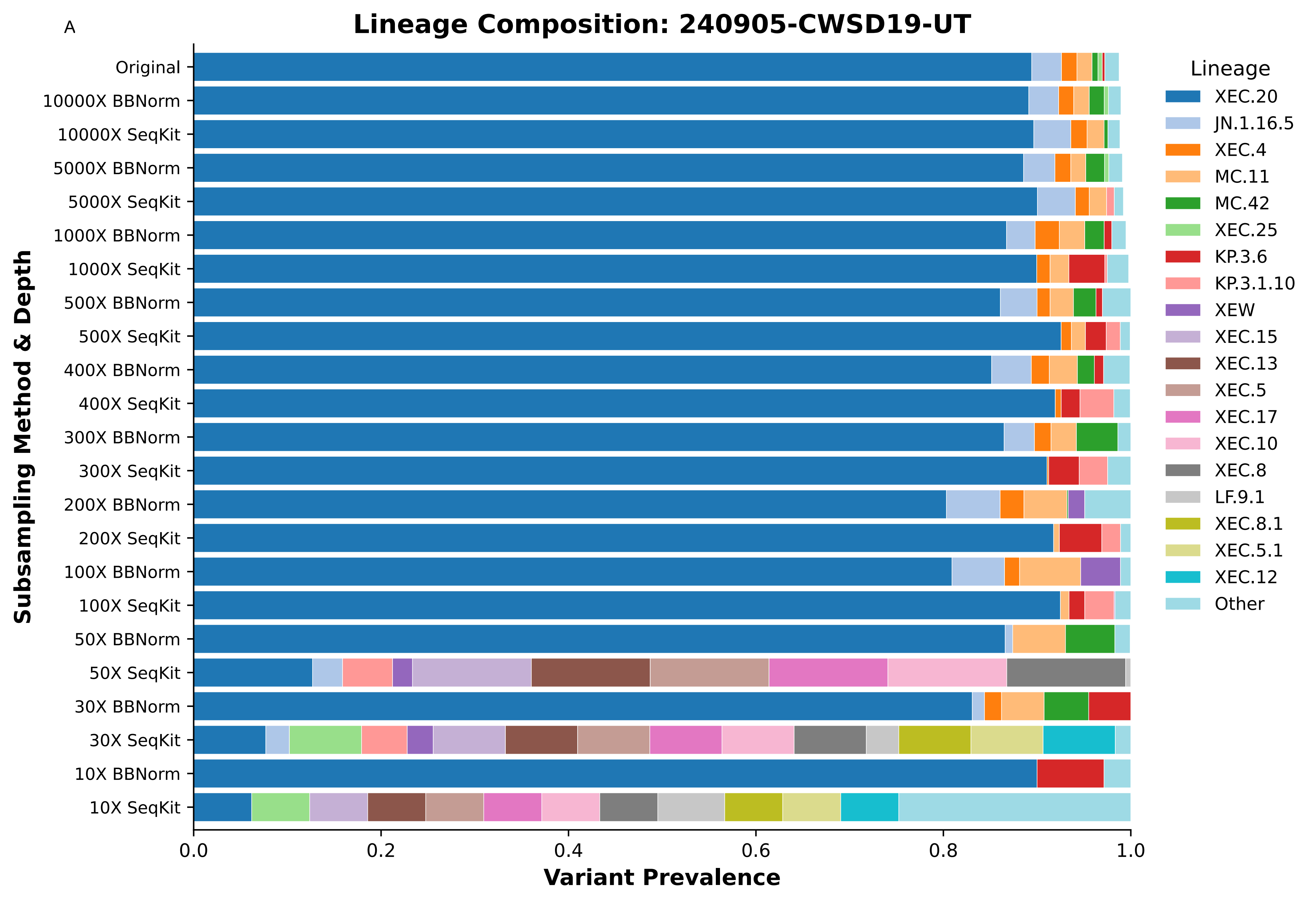
A
Lineage Composition: 240905-CWSD19-UT
Subsampling Method & Depth
Original
10000X BBNorm
10000X SeqKit
5000X BBNorm
5000X SeqKit
1000X BBNorm
1000X SeqKit
500X BBNorm
500X SeqKit
400X BBNorm
400X SeqKit
300X BBNorm
300X SeqKit
200X BBNorm
200X SeqKit
100X BBNorm
100X SeqKit
50X BBNorm
50X SeqKit
30X BBNorm
30X SeqKit
10X BBNorm
10X SeqKit
0.0
0.2
0.4
0.6
0.8
1.0
Variant Prevalence
Lineage
XEC.20
JN.1.16.5
XEC.4
MC.11
MC.42
XEC.25
KP.3.6
KP.3.1.10
XEW
XEC.15
XEC.13
XEC.5
XEC.17
XEC.10
XEC.8
LF.9.1
XEC.8.1
XEC.5.1
XEC.12
Other

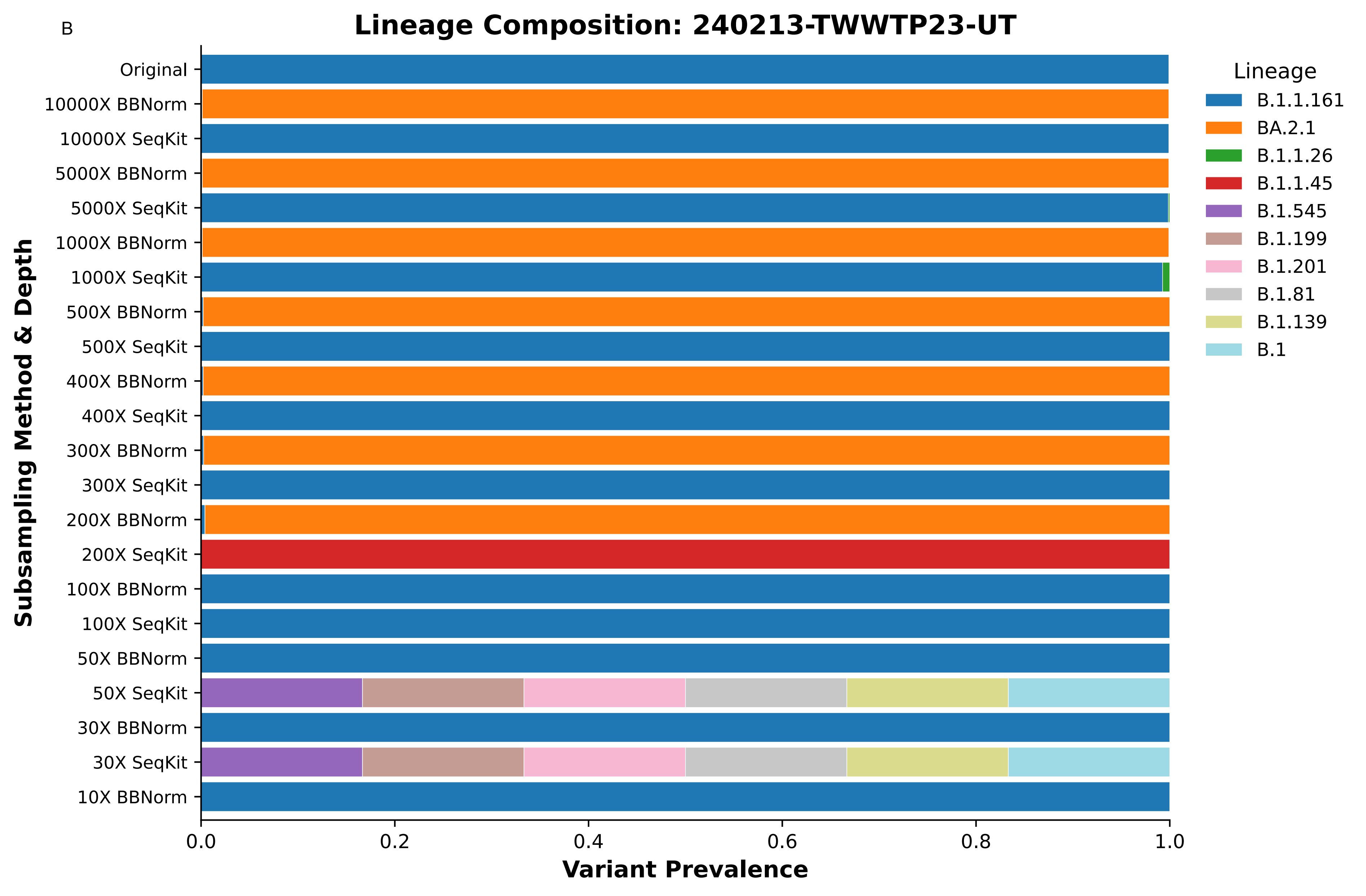

B
Lineage Composition: 240213-TWWTP23-UT
Subsampling Method & Depth
Original
10000X BBNorm
10000X SeqKit
5000X BBNorm
5000X SeqKit
1000X BBNorm
1000X SeqKit
500X BBNorm
500X SeqKit
400X BBNorm
400X SeqKit
300X BBNorm
300X SeqKit
200X BBNorm
200X SeqKit
100X BBNorm
100X SeqKit
50X BBNorm
50X SeqKit
30X BBNorm
30X SeqKit
10X BBNorm
0.0
0.2
0.4
0.6
0.8
1.0
Variant Prevalence
Lineage
B.1.1.161
BA.2.1
B.1.1.26
B.1.1.45
B.1.545
B.1.199
B.1.201
B.1.81
B.1.139
B.1

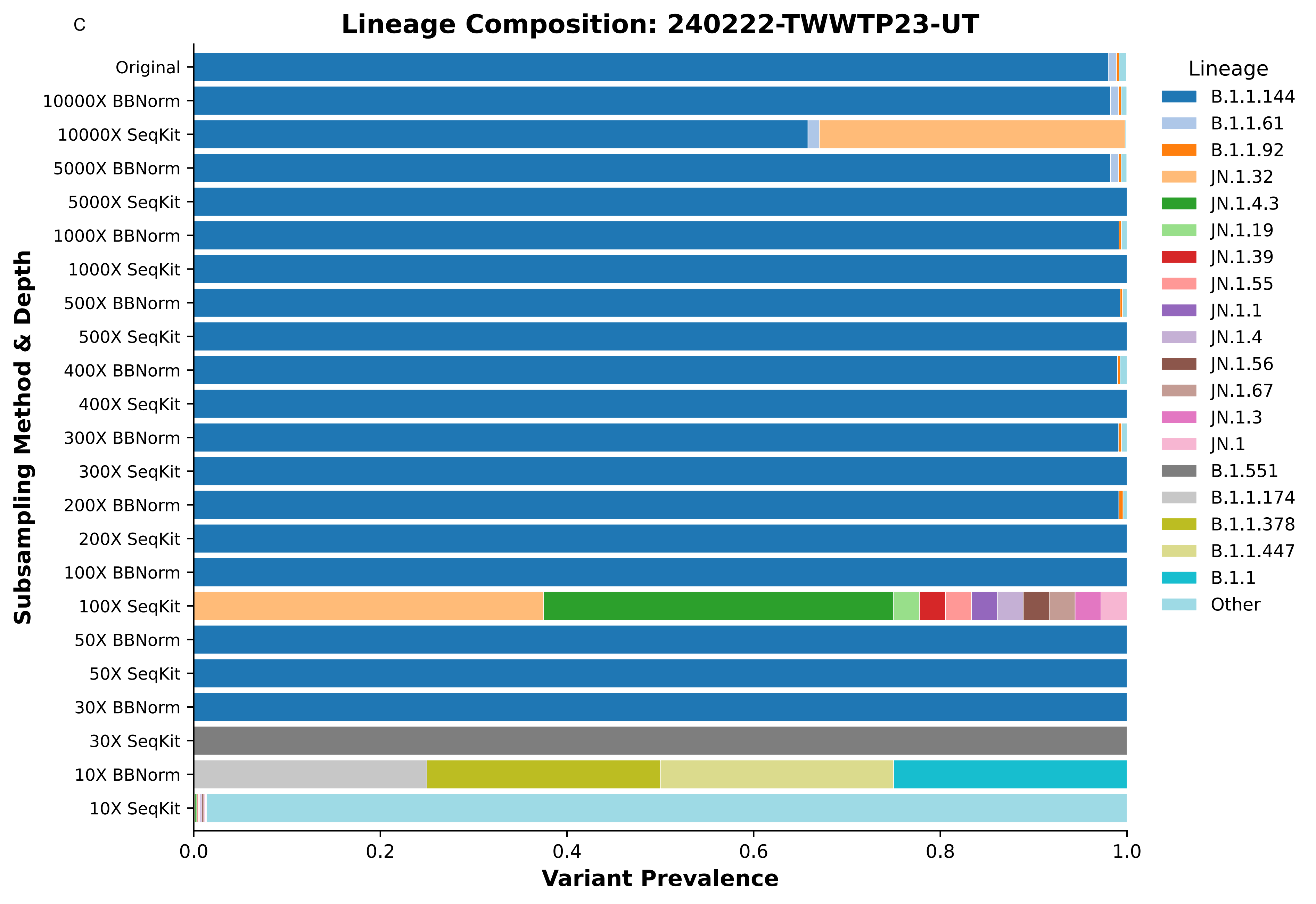
C
Lineage Composition: 240222-TWWTP23-UT
Original
10000X BBNorm
10000X SeqKit
5000X BBNorm
5000X SeqKit
1000X BBNorm
1000X SeqKit
500X BBNorm
500X SeqKit
400X BBNorm
400X SeqKit
300X BBNorm
300X SeqKit
200X BBNorm
200X SeqKit
100X BBNorm
100X SeqKit
50X BBNorm
50X SeqKit
30X BBNorm
30X SeqKit
10X BBNorm
10X SeqKit
Subsampling Method & Depth
0.0
0.2
0.4
0.6
0.8
1.0
Variant Prevalence
Lineage
B.1.1.144
B.1.1.61
B.1.1.92
JN.1.32
JN.1.4.3
JN.1.19
JN.1.39
JN.1.55
JN.1.1
JN.1.4
JN.1.56
JN.1.67
JN.1.3
JN.1
B.1.551
B.1.1.174
B.1.1.378
B.1.1.447
B.1.1
Other

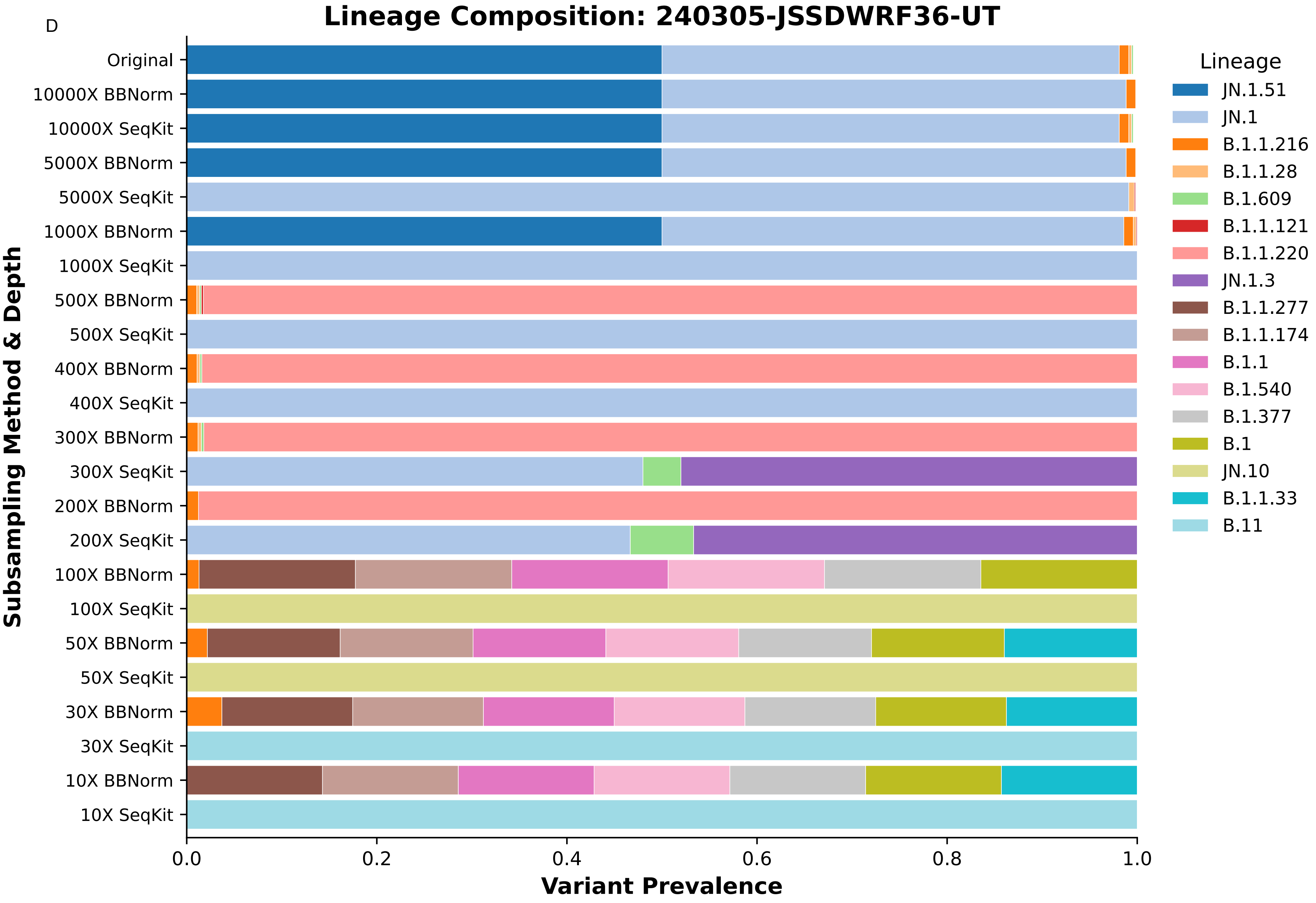

D
Lineage Composition: 240305-JSSDWRF36-UT
Subsampling Method & Depth
Original
10000X BBNorm
10000X SeqKit
5000X BBNorm
5000X SeqKit
1000X BBNorm
1000X SeqKit
500X BBNorm
500X SeqKit
400X BBNorm
400X SeqKit
300X BBNorm
300X SeqKit
200X BBNorm
200X SeqKit
100X BBNorm
100X SeqKit
50X BBNorm
50X SeqKit
30X BBNorm
30X SeqKit
10X BBNorm
10X SeqKit
Variant Prevalence
0.0
0.2
0.4
0.6
0.8
1.0
Lineage
JN.1.51
JN.1
B.1.1.216
B.1.1.28
B.1.609
B.1.1.121
B.1.1.220
JN.1.3
B.1.1.277
B.1.1.174
B.1.1
B.1.540
B.1.377
B.1
JN.10
B.1.1.33
B.11

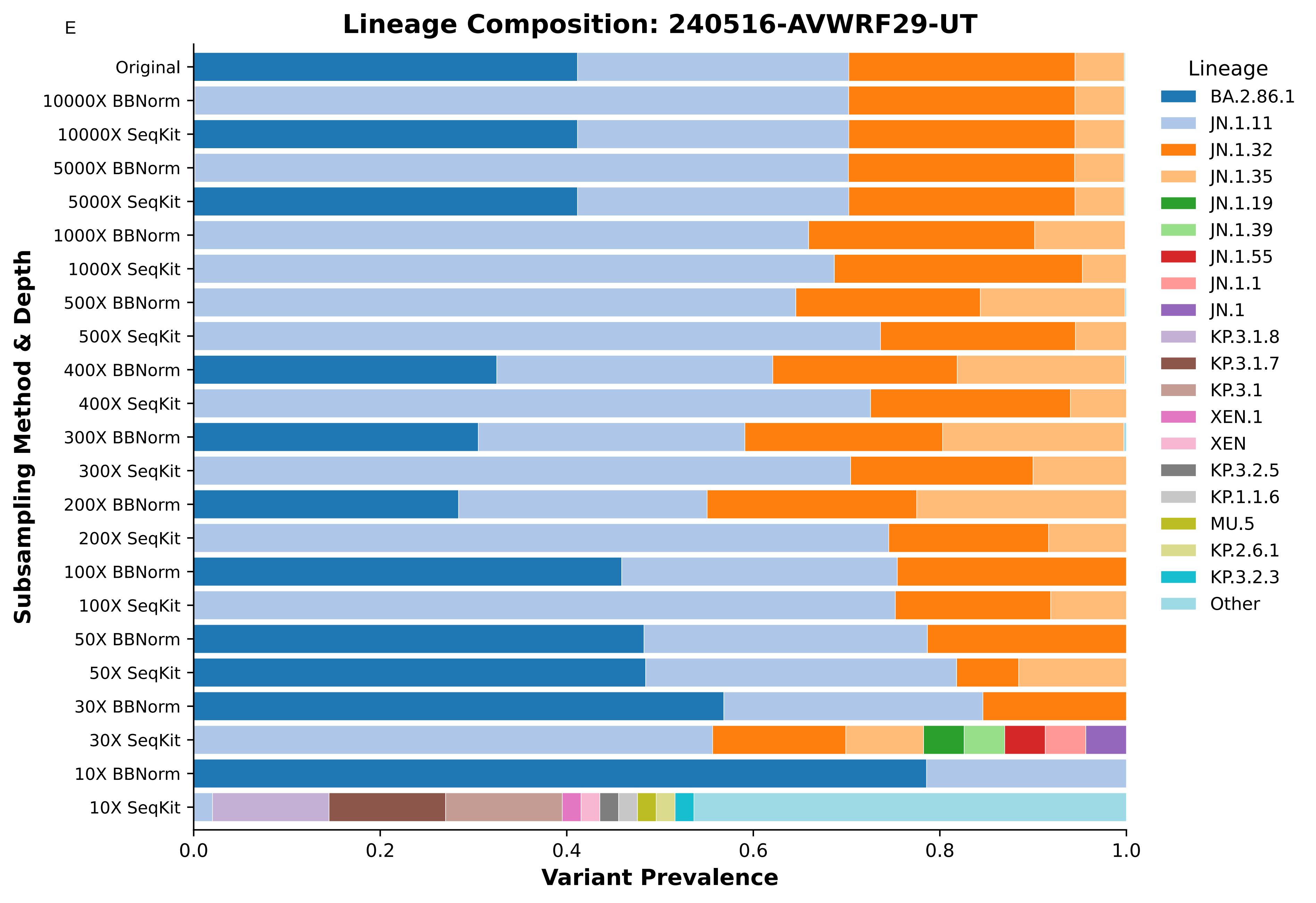
E
Lineage Composition: 240516-AVWRF29-UT
Subsampling Method & Depth
Original
10000X BBNorm
10000X SeqKit
5000X BBNorm
5000X SeqKit
1000X BBNorm
1000X SeqKit
500X BBNorm
500X SeqKit
400X BBNorm
400X SeqKit
300X BBNorm
300X SeqKit
200X BBNorm
200X SeqKit
100X BBNorm
100X SeqKit
50X BBNorm
50X SeqKit
30X BBNorm
30X SeqKit
10X BBNorm
10X SeqKit
0.0
0.2
0.4
0.6
0.8
1.0
Variant Prevalence
Lineage
BA.2.86.1
JN.1.11
JN.1.32
JN.1.35
JN.1.19
JN.1.39
JN.1.55
JN.1.1
JN.1
KP.3.1.8
KP.3.1.7
KP.3.1
XEN.1
XEN
KP.3.2.5
KP.1.1.6
MU.5
KP.2.6.1
KP.3.2.3
Other

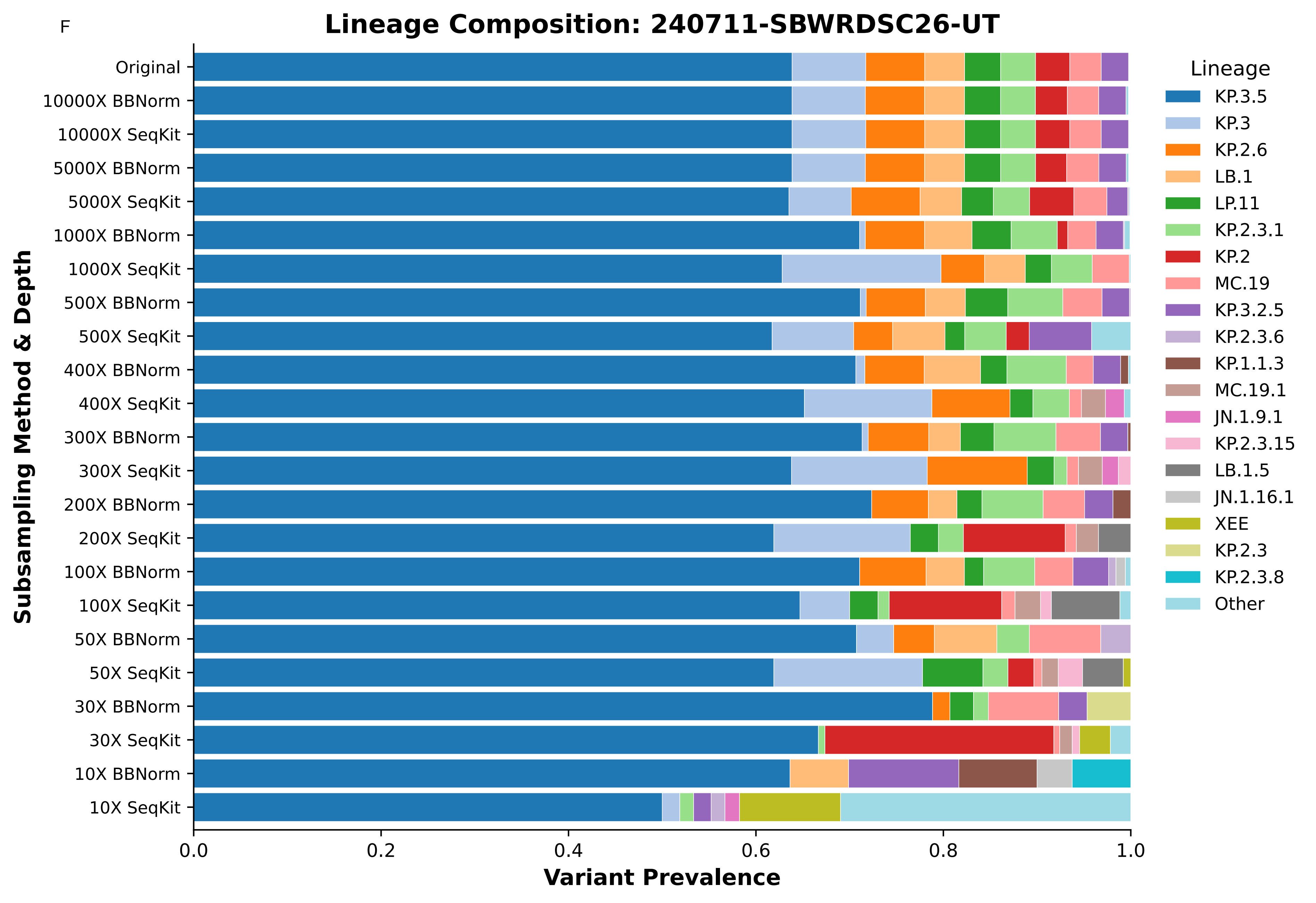

F
Lineage Composition: 240711-SBWRDSC26-UT
Subsampling Method & Depth
Original
10000X BBNorm
10000X SeqKit
5000X BBNorm
5000X SeqKit
1000X BBNorm
1000X SeqKit
500X BBNorm
500X SeqKit
400X BBNorm
400X SeqKit
300X BBNorm
300X SeqKit
200X BBNorm
200X SeqKit
100X BBNorm
100X SeqKit
50X BBNorm
50X SeqKit
30X BBNorm
30X SeqKit
10X BBNorm
10X SeqKit
0.0
0.2
0.4
0.6
0.8
1.0
Variant Prevalence
Lineage
KP.3.5
KP.3
KP.2.6
LB.1
LP.11
KP.2.3.1
KP.2
MC.19
KP.3.2.5
KP.2.3.6
KP.1.1.3
MC.19.1
JN.1.9.1
KP.2.3.15
LB.1.5
JN.1.16.1
XEE
KP.2.3
KP.2.3.8
Other

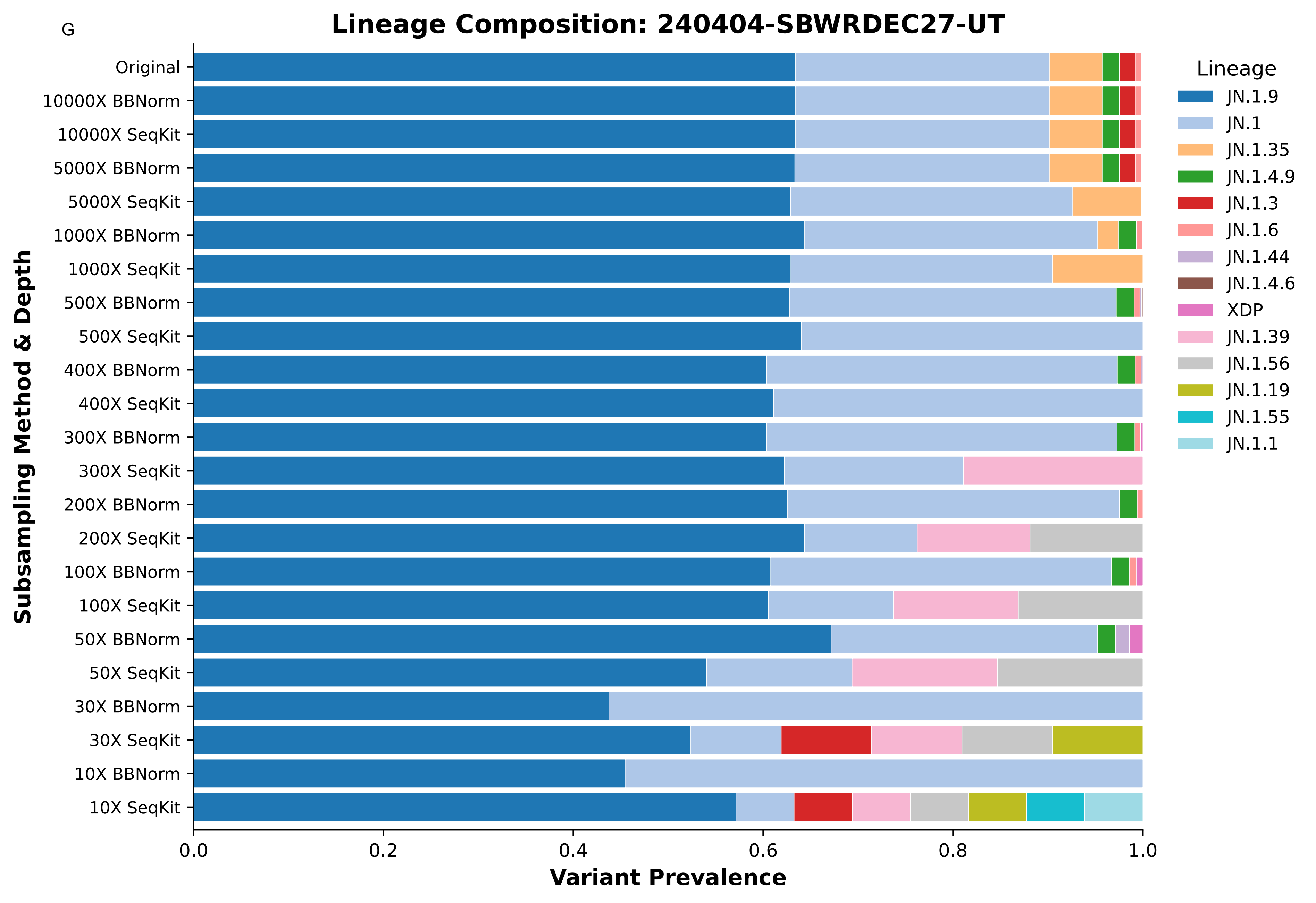
G
Lineage Composition: 240404-SBWRDEC27-UT
Subsampling Method & Depth
Original
10000X BBNorm
10000X SeqKit
5000X BBNorm
5000X SeqKit
1000X BBNorm
1000X SeqKit
500X BBNorm
500X SeqKit
400X BBNorm
400X SeqKit
300X BBNorm
300X SeqKit
200X BBNorm
200X SeqKit
100X BBNorm
100X SeqKit
50X BBNorm
50X SeqKit
30X BBNorm
30X SeqKit
10X BBNorm
10X SeqKit
0.0
0.2
0.4
0.6
0.8
1.0
Variant Prevalence
Lineage
JN.1.9
JN.1
JN.1.35
JN.1.4.9
JN.1.3
JN.1.6
JN.1.44
JN.1.4.6
XDP
JN.1.39
JN.1.56
JN.1.19
JN.1.55
JN.1.1

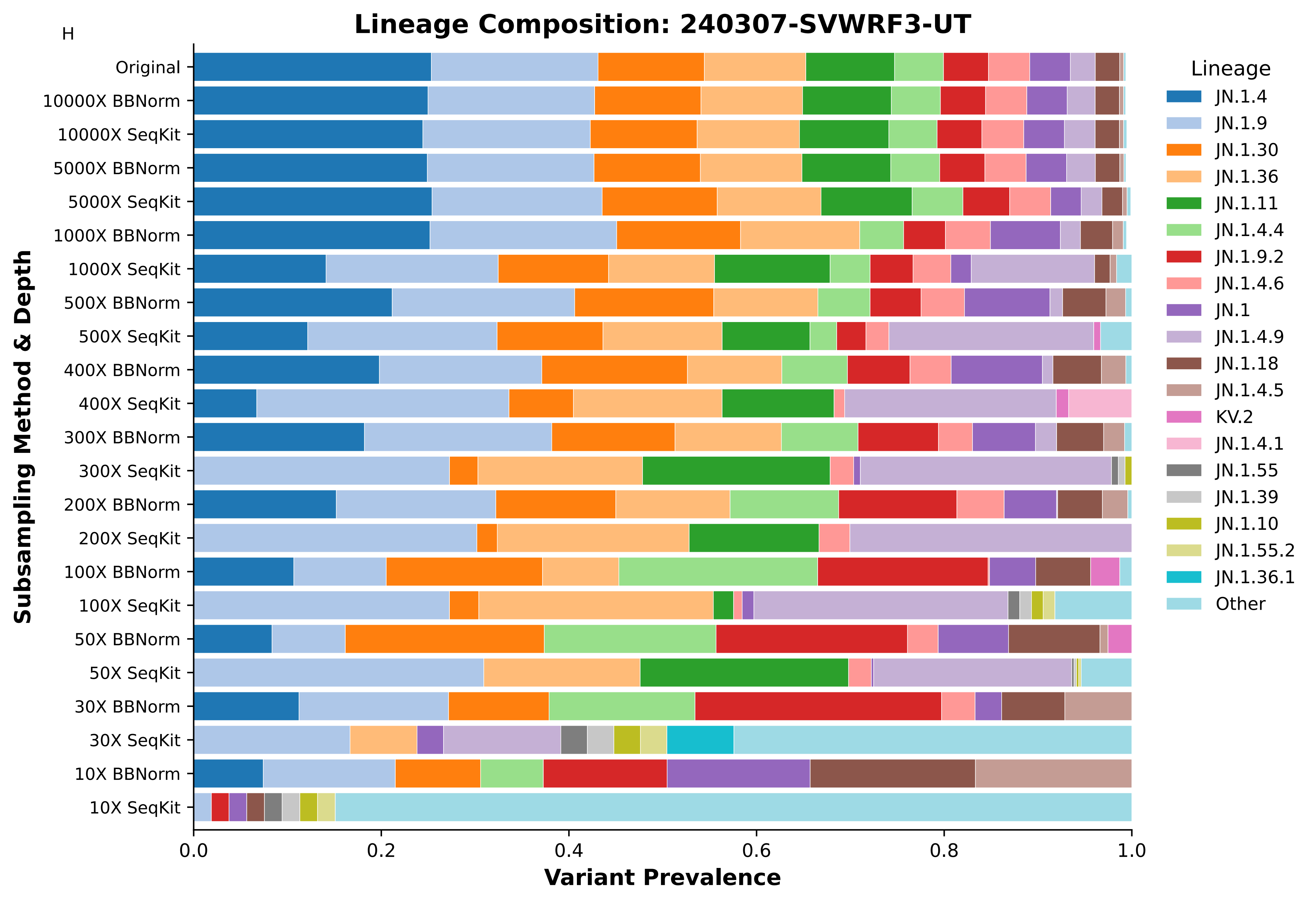
H
Lineage Composition: 240307-SVWRF3-UT
Original
10000X BBNorm
10000X SeqKit
5000X BBNorm
5000X SeqKit
1000X BBNorm
1000X SeqKit
500X BBNorm
500X SeqKit
400X BBNorm
400X SeqKit
300X BBNorm
300X SeqKit
200X BBNorm
200X SeqKit
100X BBNorm
100X SeqKit
50X BBNorm
50X SeqKit
30X BBNorm
30X SeqKit
10X BBNorm
10X SeqKit
Subsampling Method & Depth
0.0
0.2
0.4
0.6
0.8
1.0
Variant Prevalence
Lineage
JN.1.4
JN.1.9
JN.1.30
JN.1.36
JN.1.11
JN.1.4.4
JN.1.9.2
JN.1.4.6
JN.1
JN.1.4.9
JN.1.18
JN.1.4.5
KV.2
JN.1.4.1
JN.1.55
JN.1.39
JN.1.10
JN.1.55.2
JN.1.36.1
Other

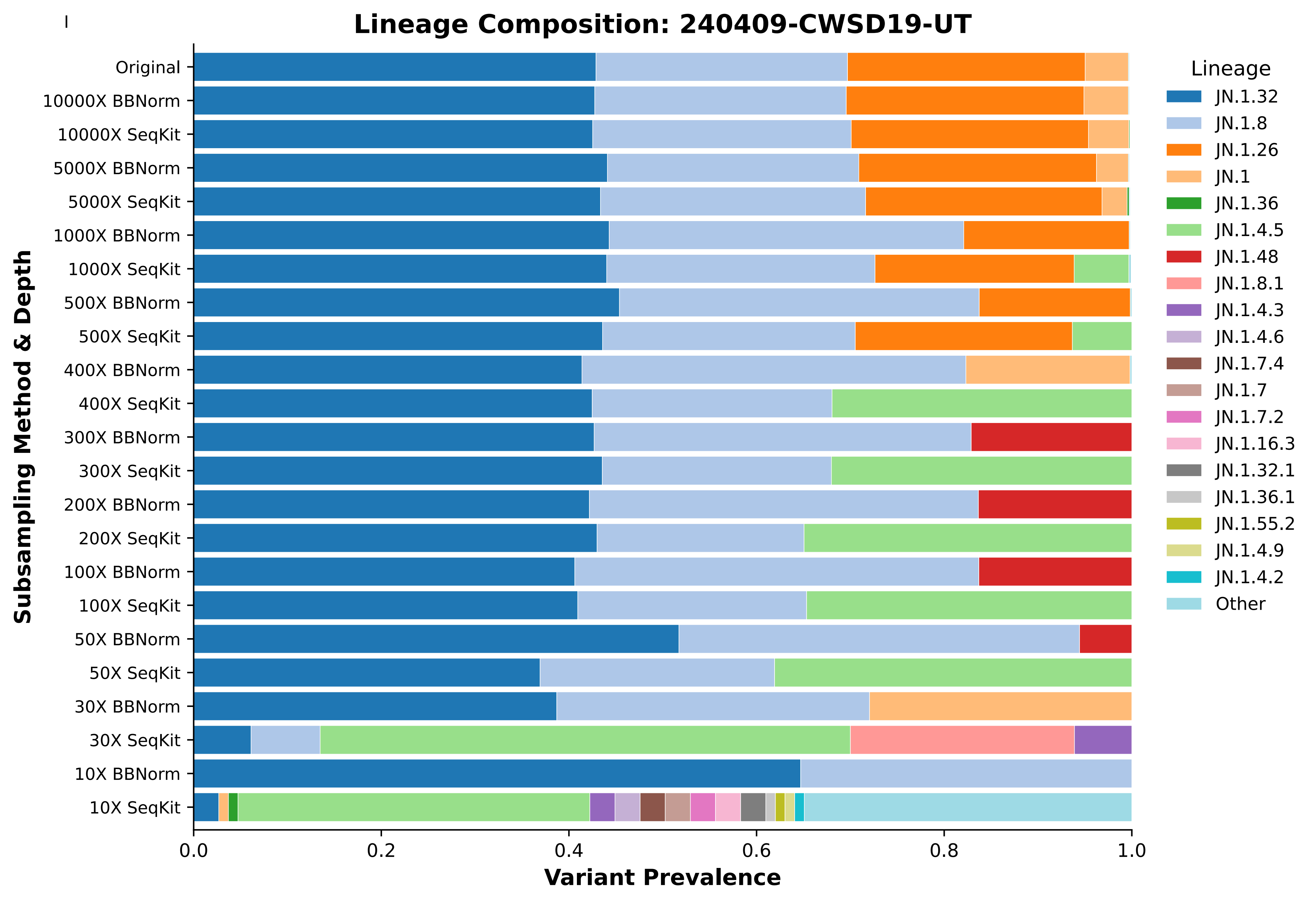
I
Lineage Composition: 240409-CWSD19-UT
Subsampling Method & Depth
Original
10000X BBNorm
10000X SeqKit
5000X BBNorm
5000X SeqKit
1000X BBNorm
1000X SeqKit
500X BBNorm
500X SeqKit
400X BBNorm
400X SeqKit
300X BBNorm
300X SeqKit
200X BBNorm
200X SeqKit
100X BBNorm
100X SeqKit
50X BBNorm
50X SeqKit
30X BBNorm
30X SeqKit
10X BBNorm
10X SeqKit
0.0
0.2
0.4
0.6
0.8
1.0
Variant Prevalence
Lineage
JN.1.32
JN.1.8
JN.1.26
JN.1
JN.1.36
JN.1.4.5
JN.1.48
JN.1.8.1
JN.1.4.3
JN.1.4.6
JN.1.7.4
JN.1.7
JN.1.7.2
JN.1.16.3
JN.1.32.1
JN.1.36.1
JN.1.55.2
JN.1.4.9
JN.1.4.2
Other

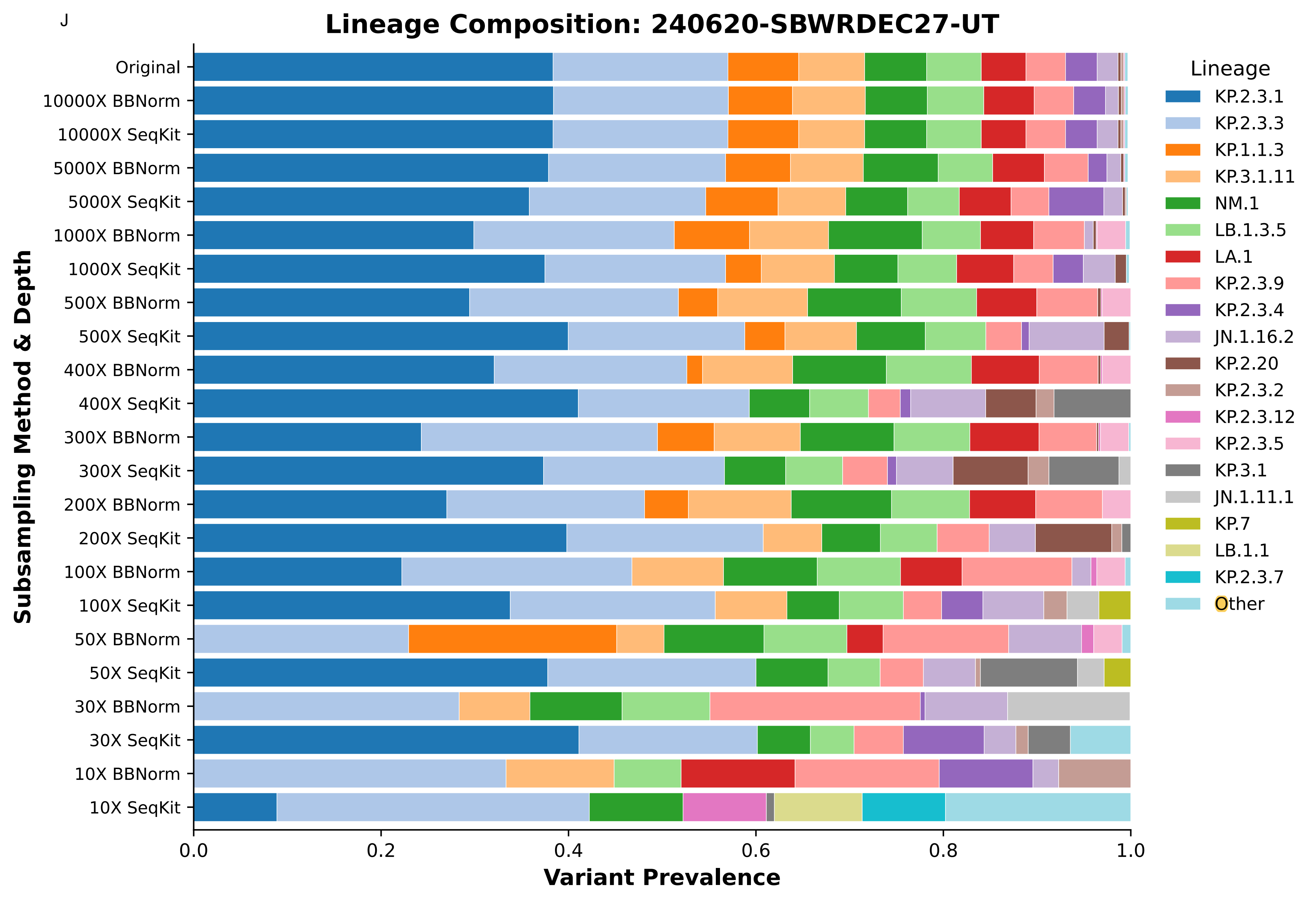
J
Lineage Composition: 240620-SBWRDEC27-UT
Subsampling Method & Depth
Original
10000X BBNorm
10000X SeqKit
5000X BBNorm
5000X SeqKit
1000X BBNorm
1000X SeqKit
500X BBNorm
500X SeqKit
400X BBNorm
400X SeqKit
300X BBNorm
300X SeqKit
200X BBNorm
200X SeqKit
100X BBNorm
100X SeqKit
50X BBNorm
50X SeqKit
30X BBNorm
30X SeqKit
10X BBNorm
10X SeqKit
0.0
0.2
0.4
0.6
0.8
1.0
Variant Prevalence
Lineage
KP.2.3.1
KP.2.3.3
KP.1.1.3
KP.3.1.11
NM.1
LB.1.3.5
LA.1
KP.2.3.9
KP.2.3.4
JN.1.16.2
KP.2.20
KP.2.3.2
KP.2.3.12
KP.2.3.5
KP.3.1
JN.1.11.1
KP.7
LB.1.1
KP.2.3.7
Other

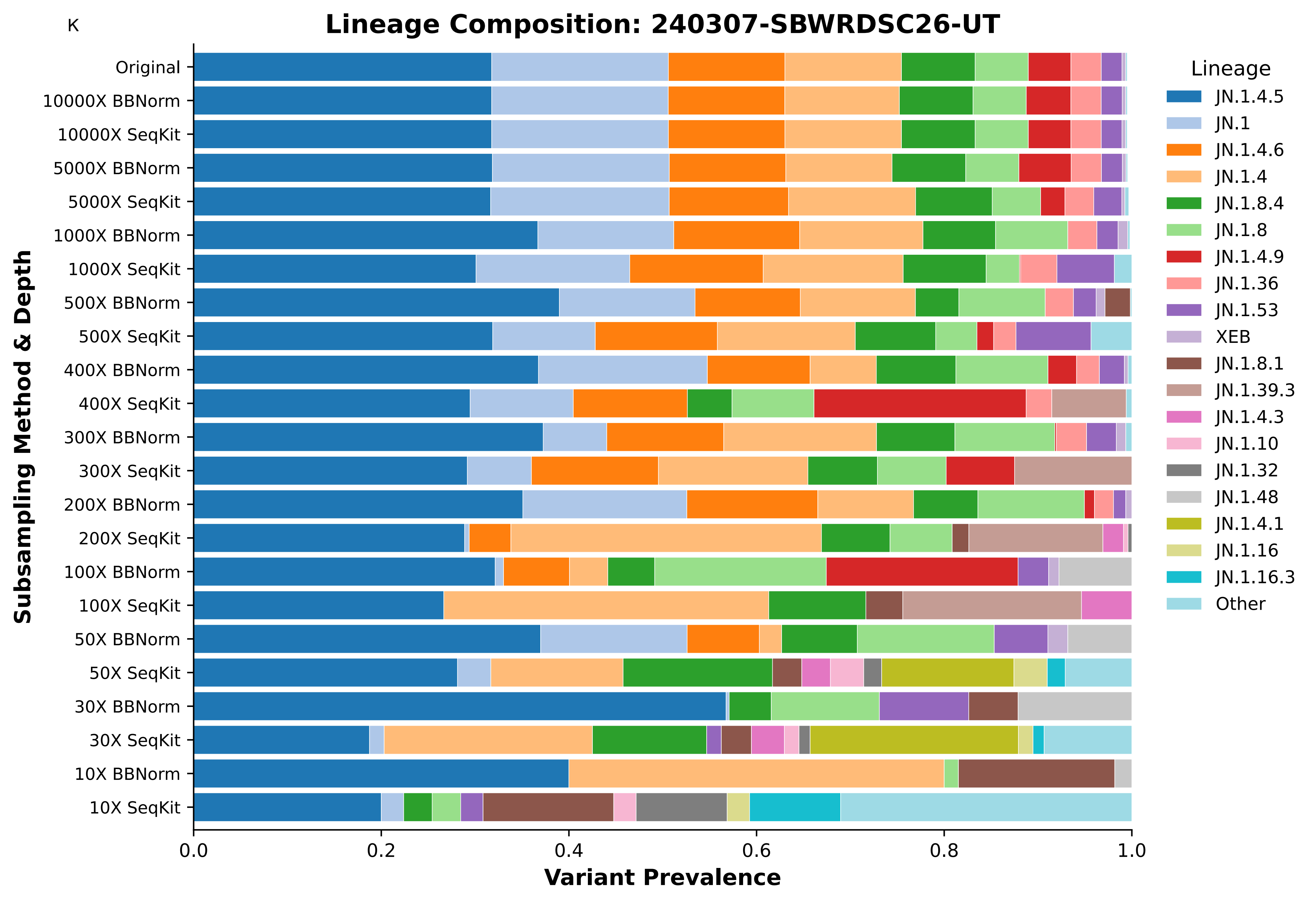
K
Lineage Composition: 240307-SBWRDSC26-UT
Subsampling Method & Depth
Original
10000X BBNorm
10000X SeqKit
5000X BBNorm
5000X SeqKit
1000X BBNorm
1000X SeqKit
500X BBNorm
500X SeqKit
400X BBNorm
400X SeqKit
300X BBNorm
300X SeqKit
200X BBNorm
200X SeqKit
100X BBNorm
100X SeqKit
50X BBNorm
50X SeqKit
30X BBNorm
30X SeqKit
10X BBNorm
10X SeqKit
0.0
0.2
0.4
0.6
0.8
1.0
Variant Prevalence
Lineage
JN.1.4.5
JN.1
JN.1.4.6
JN.1.4
JN.1.8.4
JN.1.8
JN.1.4.9
JN.1.36
JN.1.53
XEB
JN.1.8.1
JN.1.39.3
JN.1.4.3
JN.1.10
JN.1.32
JN.1.48
JN.1.4.1
JN.1.16
JN.1.16.3
Other

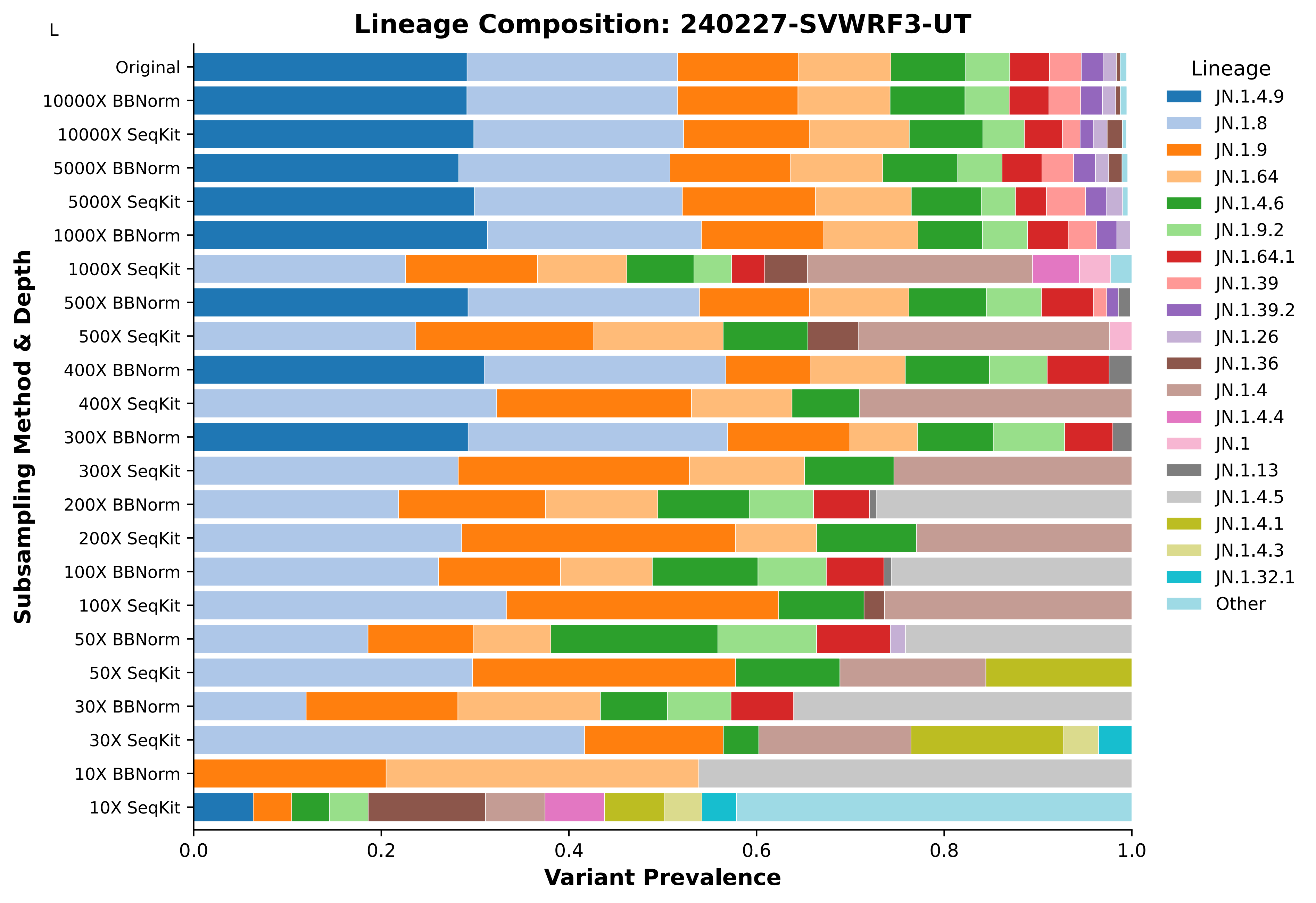

L
Lineage Composition: 240227-SVWRF3-UT
Subsampling Method & Depth
Original
10000X BBNorm
10000X SeqKit
5000X BBNorm
5000X SeqKit
1000X BBNorm
1000X SeqKit
500X BBNorm
500X SeqKit
400X BBNorm
400X SeqKit
300X BBNorm
300X SeqKit
200X BBNorm
200X SeqKit
100X BBNorm
100X SeqKit
50X BBNorm
50X SeqKit
30X BBNorm
30X SeqKit
10X BBNorm
10X SeqKit
0.0
0.2
0.4
0.6
0.8
1.0
Variant Prevalence
Lineage
JN.1.4.9
JN.1.8
JN.1.9
JN.1.64
JN.1.4.6
JN.1.9.2
JN.1.64.1
JN.1.39
JN.1.39.2
JN.1.26
JN.1.36
JN.1.4
JN.1.4.4
JN.1
JN.1.13
JN.1.4.5
JN.1.4.1
JN.1.4.3
JN.1.32.1
Other

Supplemental Figure 4. Wastewater Lineage Composition and Deconvolution Dynamics Across Target Depths. Stacked bar charts depict the estimated relative abundance of SARS-CoV-2 subvariants (deconvoluted using Freyja) across 12 individual wastewater specimens under varying data reduction conditions. Each bar compares a target read depth (10X to 10,000X) against the unprocessed 'Original' baseline for both read depth normalization (BBNorm) and random subsampling (SeqKit). Panels (A–L) correspond to individual wastewater samples (A) 240905-CWSD19-UT, (B) 240213-TWWTP23-UT, (C) 240222-TWWTP23-UT, (D) 240305-JSSDWRF36-UT, (E) 240516-AVWRF29-UT, (F) 240711-SBWRDSC26-UT, (G) 240404-SBWRDEC27-UT, (H) 240307-SVWRF3-UT, (I) 240409-CWSD19-UT, (J) 240620-SBWRDEC27-UT, (K) 240307-SBWRDSC26-UT, (L) 240227-SVWRF3-UT
