## Supplemental Figure 2 for "Data Reduction in SARS-CoV-2 Amplicon Sequencing: Effects of Read Subsampling and Normalization on Consensus Quality, Lineage Assignment, and Computational Burden"

Supplemental Figure: Per-Step Memory Usage

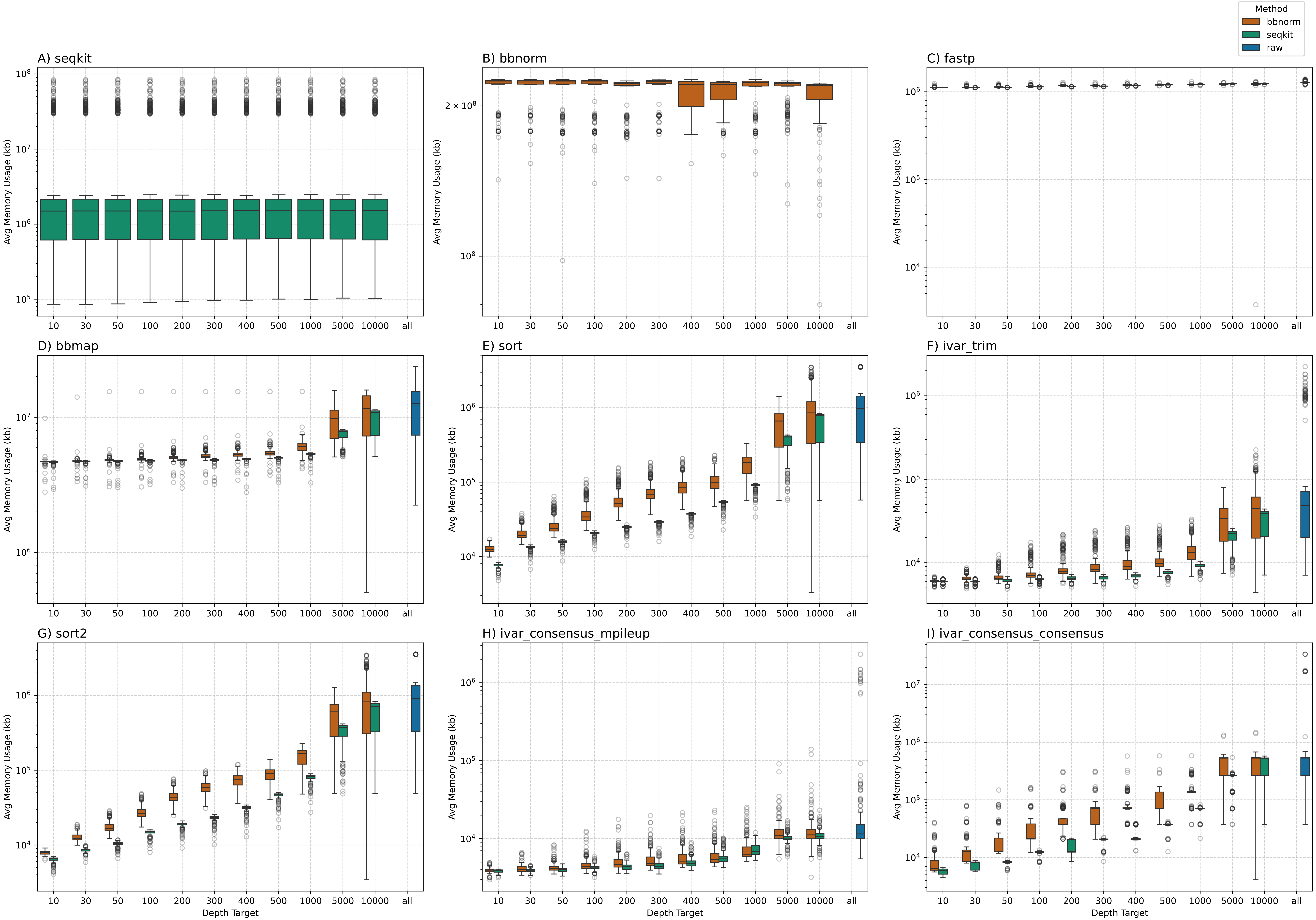

Supplemental Figure 2. Peak Memory Usage Across Individual Bioinformatic Workflow Steps. Boxplots illustrate the distribution of average peak memory usage (in kilobytes, presented on a logarithmic scale) across target read depths (10X to 10,000X and full unprocessed 'all' dataset) for each discrete step in the processing pipeline. Results compare data reduced by read depth normalization via BBNorm (orange), random subsampling via SeqKit (green), and the unprocessed baseline dataset (blue) across 220 clinical SARS-CoV-2 specimens. Individual panels depict (A) SeqKit random read subsampling, (B) BBNorm read depth normalization, (C) FASTP read quality filtering, (D) BBMap alignment to reference genome, (E) SAMtools initial alignment sorting, (F) iVar primer sequence trimming, (G) SAMtools post-trimming alignment sorting, (H) iVar mpileup generation for consensus calling, (I) iVar consensus sequence calling.
